# Combining Clinician Expertise with Prompt Engineering enhances Small Language Models Reliability for Cancer Entity Recognition in Electronic Health Records

**DOI:** 10.1101/2025.10.16.25337917

**Authors:** Federica Corso, Vittoria Peppoloni, Laura Mazzeo, Giuseppe Leone, Luana Passos, Vanja Mišković, Justin Armanini, Alberto Ferrarin, Isabella Catharina Wiest, Fabian Wolf, Giulia Montelatici, Rebecca Romanò, Ambrosini Paolo, Tommaso Capoccia, Stefano Natangelo, Simone Rota, Paola Andena, Marta De Ponti, Alessandra Russo, Giulia Stasi, Leonardo Provenzano, Andrea Spagnoletti, Marco Meazza Prina, Chiara Cavalli, Claudia Giani, Roberta Serino, Michele Borracino, Chiara Bonalume, Rosa Maria di Mauro, Claudia Agosta, Andra Diana Dumitrascu, Giorgia Di Liberti, Giulia Corrao, Teresa Beninato, Monica Ganzinelli, Mario Occhipinti, Marta Brambilla, Claudia Proto, Jakob Nicholas Kather, Alessandra Laura Giulia Pedrocchi, Filippo De Braud, Giuseppe Lo Russo, Paolo Baili, Arsela Prelaj

## Abstract

Real-world data (RWD), largely stored in unstructured electronic health records (EHRs), are critical for understanding complex diseases like cancer. However, extracting structured information from these narratives is challenging due to linguistic variability, semantic complexity, and privacy concerns. This study evaluates the performance of four locally deployable and small language models (SLMs), LLaMA, Mistral, BioMistral, and MedLLaMA, for information extraction (IE) from Italian EHRs within the APOLLO 11 trial on non-small cell lung cancer (NSCLC). We examined three prompting strategies (zero-shot, few-shot, and annotated few-shot) across English and Italian, involving clinicians with varying expertise to assess prompt design’s impact on accuracy. Results show that general-purpose models (e.g., LLaMA 3.1 8B) outperform biomedical models in most tasks, particularly in extracting binary features. Multiclass variables such as TNM staging, PD-L1, and ECOG were more difficult due to implicit language and lack of standardization. Few-shot prompting and native-language inputs significantly improved performance and reduced hallucinations. Clinical expertise enhanced consistency in annotation, particularly among students using annotated examples. The study confirms that privacy-preserving SLMs can be deployed locally for efficient and secure cancer data extraction. Findings highlight the need for hybrid systems combining SLMs with expert input and underline the importance of aligning clinical documentation practices with SLM capabilities. This is the first study to benchmark SLMs on Italian EHRs and investigate the role of clinical expertise in prompt engineering, offering valuable insights for the future integration of SLMs into real-world clinical workflows.

## 1. Introduction

Real-world data (RWD) are commonly defined as data routinely collected in hospitals and derived from electronic health records (EHRs). These data are usually stored within hospital data warehouses (DWHs), where approximately 80% of the data exist in unstructured format.^1^ Clinical narratives are crucial for understanding diseases like cancer, but manual review is slow and error-prone. Converting unstructured EHR text into structured formats could improve outcomes and research,^2,3^ though variability and complexity pose major challenges.

The emergence of large language models (LLMs) has expanded the capabilities on text classification, named entity recognition, and text summarization.^4^ LLMs are foundation models that have shown zero-shot capabilities in text generation and reasoning across a variety of NLP tasks^5^, enabling the analysis of textual data without the need for additional training or fine-tuning.^6^ In particular, the Generative Pre-Trained Transformer (GPT) by OpenAI has marked a significant turning point in generative AI with the launch of ChatGPT.^7^ Thereafter, other LLMs, including Microsoft Copilot^8^, Google Gemini 2.0^9^, Meta’s LLaMA 3^10^, Mistral AI^11^, and DeepSeek^12^ have been developed. In healthcare, two primary types of LLMs are currently being explored: general-purpose models like GPT-4, and domain-specific models such as Med-PaLM.^13^ For example, in histopathology^14^ and radiology^15^, GPT-4 has proven very good at extracting structured data from unstructured medical exams. On the other side, specialized models are trained on medical datasets and incorporate clinical guidelines, health records, and biomedical knowledge during the pre-training or fine-tuning. A growing number of researchers are evaluating LLM applications across the information extraction (IE) domain.^16,17^ Indeed, the use of EHRs poses several challenges and the context- and time-dependent semantics of clinical notes make LLM-based IE difficult and increase hallucination risk. ^18,19^ EHRs contain sensitive data, so LLM-based IE must comply with GDPR through strong de-identification and handling controls. Since many state-of-the-art models run via APIs, off-prem data transfer poses legal and ethical risks,^20^ while local deployment avoids this but often relies on lower-precision, memory-efficient quantised models.^21^

Recent studies have explored the use of small large language models (SMLs), characterized by a reduced number of parameters, typically ranging from a few million to some billion. LLaMA-7B, Gemma-7B, and Meerkat-7B offer an efficient solution for extracting medical data from clinical texts.^22,23^ These models can be deployed locally, ensuring data privacy while maintaining acceptable levels of accuracy in information extraction tasks. Finally, there is a lack of public EHRs benchmarks. To date, the MIMIC-IV^24^ is the only freely accessible EHR dataset to conduct experiments for entity recognition and relation extraction with LLMs. In this study, we benchmarked SLMs on EHRs of patients from the APOLLO 11 trial dataset^25^, a multicentric Italian study which leverages a federated model for the analysis of advanced non-small cell lung cancer (NSCLC) data. Immunotherapy (IO) targeting PD-L1 expression and Tyrosine kinase inhibitors (TKIs) for patients with targetable mutations have transformed NSCLC treatment.^26,27^ Key clinical features are central to the treatment decisions, but they are often embedded in EHRs, posing the need to leverage SLMs for automated IE. To the best of our knowledge, this is the first study benchmarking LLMs on clinical feature extraction using Italian EHRs and involving doctors into prompting experiments to investigate whether clinical expertise influences language models performance on the extraction task. In this work, we benchmarked SLMs on a real-world dataset of 897 NSCLC patients divided by development set (the IO cohort) and validation set (the TKI cohort). Four locally deployable SLMs, 2 generalist and 2 biomedical-specialized, such as LLaMA 3.1 8B, Mistral 7B, MedLLaMA 2 7B, and BioMistral 7B have been tested for IE of cancer variables. Furthermore, we tested three prompt designs, such as zero/few-shot prompting and few-shot prompting with annotated data on 11 clinical features between two different prompt languages (Italian and English). Finally, we investigated the influence of doctors’ expertise in the extraction accuracy, conducting a prompt engineering study with experienced, resident oncologists and medical students.

## 2. Material and Methods

### 2.1 Study cohorts

EHRs were extracted from the institutional data warehouse (DWH) of Fondazione IRCSS Istituto Nazionale dei Tumori of Milan. Textual data sources are Electronic Health Records 1 (EHR1) and Electronic Health Records 2 (EHR2), both stored in the DWH. The full study cohort is part of the APOLLO 11 trial^25^ (NCT05550961), in which two cohorts have been identified by cancer treatment (Figure S1.1). Written informed consent was obtained from all living participants included in the study, and in accordance with Italian legislation (GDPR EU 2016/679 and national privacy regulations), informed consent is not required for deceased individuals; their data were collected and used for research purposes under the applicable legal provisions. Patient data were collected in accordance with the Declaration of Helsinki, Good Clinical Practice and local ethical rules. The study protocol INT 128/22 was approved by the appropriate institutional ethics committee, Fondazione IRCCS Istituto Nazionale dei Tumori di Milano. The IO cohort represents the development cohort on which the LLM pipeline was first implemented. This cohort includes 711 patients treated with: i) IO in maintenance after chemo-radiotherapy, ii) first-line (1L) IO, iii) IO in subsequent lines. For these patients the following inclusion criteria have been applied: group A are patients with EHR1 within ± 7 days from the IO start date; group B are patients with an EHR1 at any date from IO start date; group C includes patients with a EHR2 at any date from IO start date. A second cohort, named the TKI cohort, is the validation cohort, on which the extraction pipeline was then validated. The TKI cohort included 186 EGFR-mutated patients treated with TKIs in 1L and subsequent lines. In this cohort, patients have been distinguished as follows: patients with an EHR1 at TKI start date after 2018 and patients with an EHR from EHR2 at TKI start date before 2018. Texts were pseudonymized and pre-processed as reported in Section 2.3.

### 2.2 Data pre-processing and anonymization

In the pre-processing phase, we implemented only essential text cleaning operations, specifically HTML tag removal to address potential web scraping artifacts and expansion of common Italian contractions (e.g., “dr.ssa” → “dottoressa”) to maintain text homogeneity. The anonymization process has been designed as a two-stage pipeline: entity identification and masking operations. The identification phase was implemented as a NER task. We utilized the Presidio Python package (from Microsoft)^28^ with the spaCy “it_core_news_lg” Italian model to identify mentions of people (“PERSON” entities). We used pattern-based recognition for Italian-specific identifiers such as “IT_FISCAL_CODE,” “IT_IDENTITY_CARD,” “EMAIL_ADDRESS,” and “PHONE_NUMBER.” To mitigate false positive classifications arising from ambiguous contexts, such as eponymous diseases incorporating discoverer names (e.g., “Linfoma di Hodgkin”), we relied on domain-specific lookup tables containing curated lists of medical conditions and pharmaceutical compounds. These two tables are fundible within the DWH. For the masking phase, we employed entity type substitution, replacing identified sensitive information with corresponding entity placeholders (e.g., <PERSON>, <EMAIL_ADDRESS>). This approach preserves the semantic and syntactic context essential for LLM comprehension^29^ while maintaining privacy protection, as these models leverage contextual relationships to inform downstream task performance.

### 2.3 Entity recognition and prompt design

For clinical entity recognition, we adopted LLM-AIx^30^, an open-source pipeline for IE that works via an easy-to-use interface leveraging privacy-preserving LLMs. The protocol consists of four main processing steps: 1) problem definition and data preparation, 2) data preprocessing, 3) LLM-based IE and 4) output evaluation (Figure 1). In particular, the LLM-based IE step allows user specifications on the model to run and the prompt definition. Model configuration includes temperature and maximum number of generated tokens. In our prompts, we first introduce the context and then we instruct the model with the task. For more complex features, the prompt is eventually enriched with examples in the same format of the output. Three prompting strategies have been tested: 1) zero-shot prompting directly asks the model to extract clinical feature without input/output examples, 2) few-shot prompting provides the model with a few generic input/output examples, 3) few-shot prompting with annotated data (few shot + ann.)^31^ enriches the prompt with annotated examples from our specific dataset. To ensure consistent output the LLM is forced to follow a JSON schema through a grammar builder as the one presented in Figure S2.1 that helps the user to assign a label to the feature to be extracted and to select the desired output format (e.g., boolean, multiclass). For further details, we refer to the original publication of Wiest et al.^32^ Extraction from EHRs was assessed for the following 11 clinical features (Table S2.2): smoking (current/former/never), ECOG PS (0-3), histology (adenocarcinoma/squamous cell carcinoma/other histologies), PD-L1 (<1; 1-49%; ≥50%), overall staging (III/IV), primary tumor features (T0-T4), regional lymph node involvement (N0-N3), and distant metastases (M0/M1), as multiclass variables and bone/brain/liver metastases as boolean variables (TRUE/FALSE). One of the major concerns for LLMs is the risk of hallucinations when the extraction is not supported by any evidence in the text. For these cases, we assigned a “not mentioned” category to 8 features except for the 3 metastasis sites for which no mention has been treated as absence of metastasis. Finally, we used the REDCap electronic Case Report Form for APOLLO11 as ground truth (GT) with part of the data manually revised by our in-training oncologists and data scientists for further annotations.

**Figure 1:**
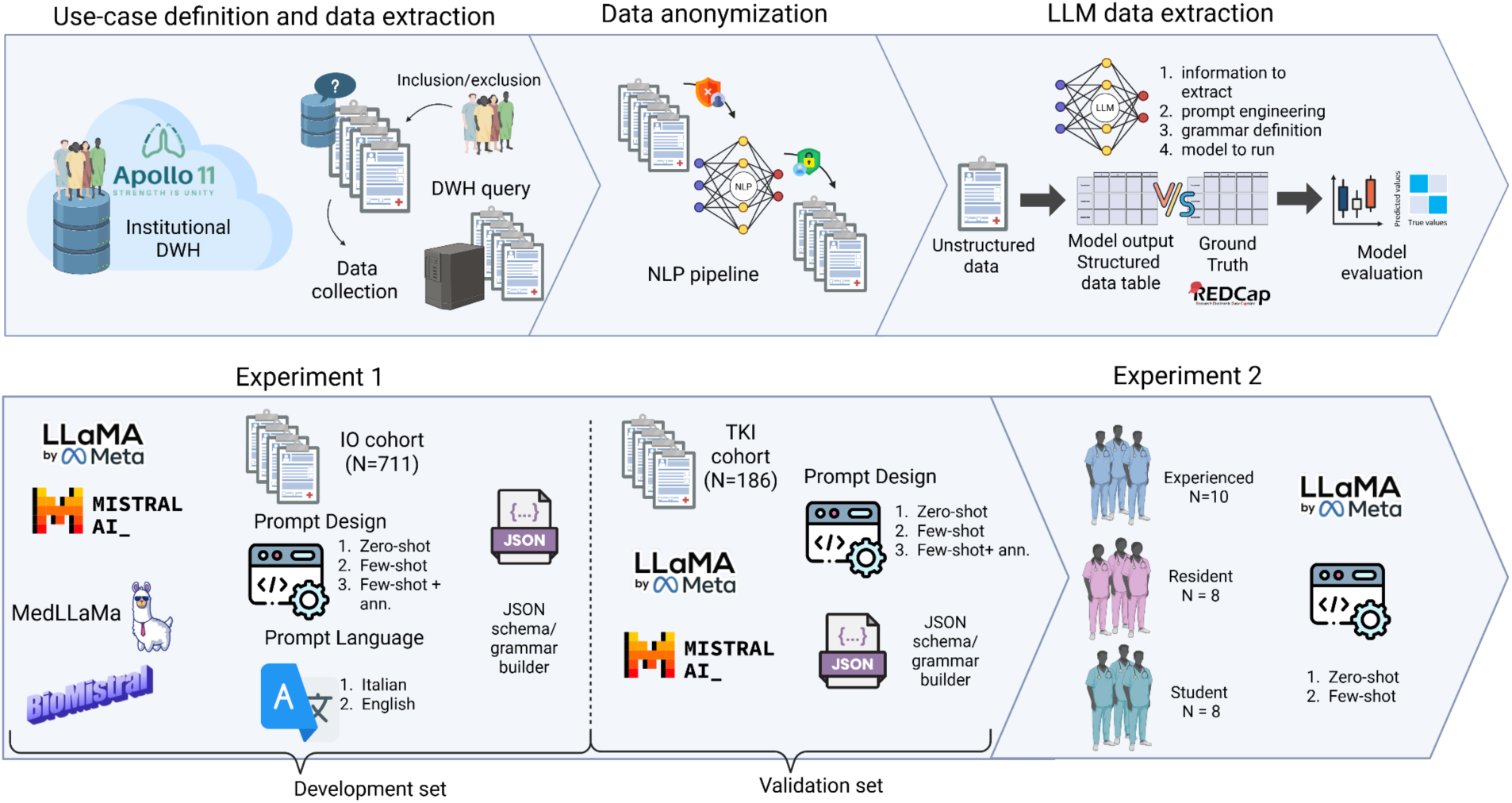
Overview of the experimental pipeline for LLM-based data extraction. The process includes data retrieval from the institutional DWH, anonymization via NLP pipelines, and structured data extraction using LLMs. Experiment 1 evaluates multiple models, prompt types, and languages across two clinical cohorts. Experiment 2 assesses the impact of clinical expertise (experienced, resident, student) on zero-shot and few-shot prompt design using LLaMa 3.1.

### 2.4 Quantized Large Language models

In this study, we evaluated the performance of four state-of-the-art, open-source LLMs for clinical information extraction: LLaMA 3.1 8B, Mistral 7B, MedLLaMA 2 7B, and BioMistral 7B. These models differ in both scale and domain specialization. While LLaMA and Mistral represent general-purpose LLMs, MedLLaMA and BioMistral introduce domain alignment through additional biomedical pre-training. This diversity allows us to evaluate how domain-specific adaptation influences extraction accuracy in unstructured clinical narratives. All models were utilized in their pre-quantized formats to ensure computational efficiency and memory optimization. We employed LLaMA 3.1 8B Instruct in 8-bit quantization, MedLLaMA 2 7B in 5-bit quantization, Mistral 7B Instruct v0.3 in 4-bit quantization and BioMistral 7B in 8-bit quantization. These quantization levels represent the best trade-off between model compression and performance retention.

### 2.5 Clinical expertise

A total of 26 medical experts with different levels of experience have been involved in this experiment: 10 experienced oncologists (> 5 years of practice), 8 resident oncologists (1-4 years of practice), 8 medical students (last year of university). To assess whether clinical experience impacts on the LLM extraction accuracy, we conducted a prompt engineering experiment. In phase 1, participants were asked to write zero-shot prompts for extracting clinical information from EHRs. In phase 2, participants refined their prompts with few-shot examples according to results obtained in phase 1. A prompt template has been submitted to medical doctors to ensure consistency among participants. The doctors were asked to prompt the model for the following features: T, N, M, overall staging and bone/brain/liver metastases. Only LLaMA 3.1 8B on the IO dataset was used for this analysis.

### 2.6 Model evaluation and statistical analysis

We estimated accuracy, precision, recall, and F1-Score with 95% binomial CIs to evaluate model performance against the human-annotated GT. To test whether LLM correctly captures when the feature is missing (i.e., the “not mentioned” label) in the text, we implemented an error analysis specific to missing information (MI) retrieval. For each feature, we collected as percentages the true missing information and the predicted missing information (i.e., hallucination rate). This analysis has been conducted for zero and few-shot + ann. prompting strategies. Inter-model agreement was measured with Fleiss κ statistics.^33^ Fleiss κ values are interpreted as follows: less than 0 = poor, 0–0.20 = slight, 0.21–0.40 = fair, 0.410.60 = moderate, 0.61–0.80 = substantial, and 0.81–1.00 = almost perfect. Wilcoxon signed-rank test for paired samples was used to compare agreement between two sets of methods and false discovery rate^34^ was used to correct for multiple comparisons. Performance between prompts in Italian and English was tested with a McNemar (for binary) and Bowkey (for multiclass) test. Finally, clinical expertise analysis was carried out comparing, for each feature, the intra and inter-rater Fleiss κ agreement^33^ (e.g., experts and experts vs students) between the zero and few-shot phases of the experiment using the Wilcoxon rank test. P < .05 was considered for statistical significance. Statistical analyses were performed using Python version 3.12 using the library SciPy (1.16.0).

## 3. Results

### 3.1 IO and TKI patient characteristics

Clinical and pathological characteristics of the IO and TKI cohorts are summarized in Table 1. IO cohort includes 711 patients with adenocarcinoma as the predominant histological subtype (66.4%), followed by squamous cell carcinoma (18.9%). Most patients had a history of smoking (43.2%), and PD-L1 positivity (≥1%) was observed in 51.5% of the cases. ECOG PS was available in the clinical reports for most patients, with 49.8% of patients scoring 1 and 12.1% scoring 2. Bone metastases were present in 39.9% of patients, while liver and brain metastases were noted in 17.3% and 19.8%, respectively. Regarding TNM staging, 46.3% were classified as T4, and 90.3% were staged as M1. The TKI cohort includes 186 patients with adenocarcinoma (95.16%) as the most common subtype and PDL1 <1% in most patients. As in the IO cohort, majority of the patients were scored with ECOG PS 0-1. Bone and brain metastases were present in 50.54% and 31.18% of patients, respectively. Finally, TNM staging had missing information in the majority of reports as well as overall staging (68.28%).

**Table 1:** Patient characteristics of the APOLLO 11 dataset divided by IO and TKI cohorts.

| Characteristics | Development set | Validation set |
| --- | --- | --- |
|  | IO cohort (n=711) | TKI cohort (n=186) |
| **Sex** |  |  |
| Male | 449 (63%) | 65 (34.94%) |
| Female | 262 (37%) | 123 (66.12%) |
| **Age** |  |  |
| Median (Q1-Q3) | 68 (61-74) | 67.5 (58-73) |
| **Histological subtype** |  |  |
| Adenocarcinoma | 472 (66.4%) | 177 (95.16%) |
| Squamous cell carcinoma | 134 (18.9%) | 7 (3.76%) |
| Other histology | 99 (13.9%) | 2 (1.08%) |
| Not mentioned | 6 (0.8%) | 0 (0%) |
| **Smoking history, n (%)** |  |  |
| Current smoker | 127 (17.9%) | 18 (9.68%) |
| Former smoker | 307 (43.2%) | 58 (31.18%) |
| Never smoked | 42 (5.9%) | 42 (22.58%) |
| Not mentioned | 235 (33.0%) | 68 (36.56%) |
| **PD-L1 expression** |  |  |
| <1% | 145 (20.4%) | 51 (27.42%) |
| 1–49% | 192 (27.0%) | 35 (18.82%) |
| > 50% | 174 (24.5%) | 23 (12.37%) |
| Not mentioned | 200 (28.1%) | 77 (41.4%) |
| **ECOG PS** |  |  |
| 0 | 194 (27.3%) | 23 (12.37%) |
| 1 | 354 (49.8%) | 50 (26.88%) |
| 2 | 86 (12.1%) | 37 (19.89%) |
| 3 | 10 (1.4%) | 22 (11.83%) |
| 4 | 0 (0%) | 2 (1.08%) |
| Not mentioned | 67 (9.4%) | 52 (27.96%) |
| **Presence of metastasis** |  |  |
| Bone | 284 (39.9%) | 94 (50.54%) |
| Liver | 123 (17.3%) | 20 (10.75%) |
| Brain | 141 (19.8%) | 58 (31.18%) |
| **T stage** |  |  |
| T0 | 90 (12.7%) | 0 (0%) |
| T1 | 52 (7.3%) | 5 (2.69%) |
| T2 | 87 (12.2%) | 18 (9.68%) |
| T3 | 148 (20.8%) | 14 (7.53%) |
| T4 | 329 (46.3%) | 16 (8.60%) |
| Not mentioned | 5 (0.7%) | 133 (71.51%) |
| **N stage** |  |  |
| N0 | 105 (14.8%) | 105 (14.8%) |
| N1 | 50 (7.0%) | 2 (1.08%) |
| N2 | 334 (47.0%) | 17 (9.14%) |
| N3 | 218 (30.7%) | 31 (16.67%) |
| Not mentioned | 4 (0.6%) | 136 (73.12%) |
| **M stage** |  |  |
| M0 | 69 (9.7%) | 4 (2.15%) |
| M1 | 642 (90.3%) | 107 (57.53%) |
| Not mentioned | 0 (0%) | 75 (40.32%) |
| **Stage** |  |  |
| III | 63 (8.9%) | 3 (1.61%) |
| IV | 643 (90.4%) | 56 (30.11%) |
| Not mentioned | 5 (0.7%) | 127 (68.28%) |

### 3.2 IO cohort results

#### 3.2.1 Benchmark of models and prompt design

We compared the performance of four privacy-preserving LLMs using three different prompting techniques (zero-shot, few-shot and few-shot+ ann) across 11 clinical variables. Figure 2 presents the heatmap of model performance for both accuracy and F1-score for all the features using all prompting techniques. LlaMa 3.1 achieved the highest average accuracy (0.90), followed by Mistral 7B (0.69), BioMistral 7B (0.44), and MedLlama 2 (0.27). For ECOG PS zero-shot accuracy is high (LlaMa 3.1 0.93 [0.91, 0.95], Mistral 0.89 [0.86, 0.91]), and it reaches ≥ 94 % with few-shot and annotation-based prompting (with LLaMa in Few-shot + clinical ann. 0.97 [0.95, 0.98]. Histology and brain metastasis also show strong zero-shot performance (LlaMa 3.1: 0.87 [0.84, 0.89] and 0.91 [0.88, 0.93]; Mistral: 0.85 [0.82, 0.88] and 0.86 [0.83, 0.88]) and improve further with annotated examples. For smoking, LlaMa 3.1 zero-shot is 0.72 [0.69, 0.75] and improves to 0.87 [0.84, 0.89] with annotation, whereas Mistral never exceeds 0.67 [0.64, 0.70]. PD-L1 expression shows moderate zero-shot accuracy (LlaMa 3.1 0.76 [0.73, 0.79], Mistral 0.81 [0.78, 0.84]) and modest improvements with few-shot and annotation. Metastatic sites extraction (bone, liver, brain) showed consistently high accuracy across top-performing models, with LlaMa 3.1 achieving > 0.90 accuracy for all sites. T, N, M staging are the features performing the worst. For both models, T stage extraction ranges around 20–50 % accuracy across all prompting strategies (LlaMa 3.1 zero-shot 0.47 [0.43, 0.50], few-shot 0.39 [0.36, 0.43], annotated 0.48 [0.45, 0.52]; Mistral zero-shot 0.24 [0.21, 0.27], few-shot 0.23 [0.20, 0.25], annotated 0.21 [0.18, 0.24]). N stage is slightly better (around 45–55 %); M stage and overall stage show better prediction (LlaMa 3.1 few-shot/annotated M ≈ 91–92 %, Stage ≈ 90–91 %; Mistral few-shot M ≈ 65–73 %, Stage ≈ 84–85 %). Similar patterns were observed for F1-scores, with LlaMa 3.1 demonstrating superior average performance (Macro F1 score: 0.62), followed by Mistral (Macro F1 score: 0.54), BioMistral (Macro F1 score: 0.29), and MedLlama (Macro F1 score: 0.24). In contrast with high accuracy, the lower F1-score of overall staging and histology reflect the unbalanced nature of these features. In addition, we selected 54 pathology reports from the IO patients who underwent a surgery and for which pathological T (pT) and N (pN) was evaluated. On average, this analysis returned perfect accuracy and F1-score for pT and accuracy and F1-score equal to 0.98 for pN. No differences in performance were observed across prompt design. Details of LlaMa 3.1 and Mistral performance with Italian prompts in the IO cohort are reported in Tables S3.1and S3.2.

**Figure 2:**
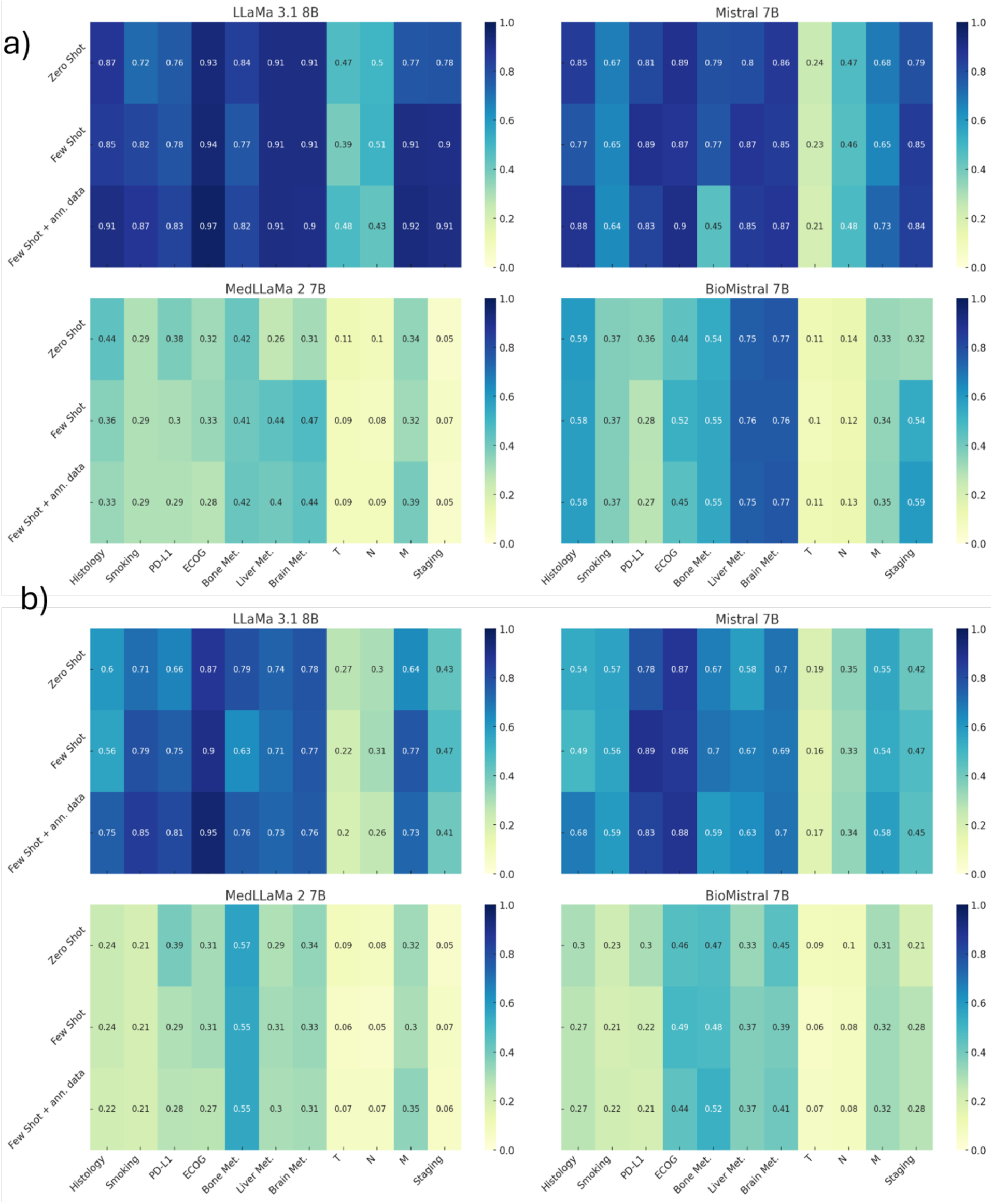
heatmaps of accuracy a) and b) F1-score for LLaMa 3.1 8B, Mistral 7B, MedLLaMa 2 7B and BioMistral 7B by prompting techniques on the IO cohort.

#### 3.2.2 Prompt language comparison

In a second analysis, we assessed whether prompt language significantly affects the model’s ability to extract structured cancer information. We investigated for each prompting design the impact of both English and Italian prompts with LLaMA 3.1 (Tables S4.1 and S4.2). Overall, prompting in Italian led to significantly better extraction performance for several features (Figure 3), most notably smoking history, where accuracy improved from 48% (English) to 72% (Italian) in the zero-shot setting (p < 0.01), and from 59% to 82% in the few-shot setting (p < 0.01). Similarly, for bone metastases, Italian prompts outperformed English in few-shot prompting with an F1-score increase from 0.31 to 0.63 (p < 0.01). In contrast, for features like PD-L1, performance favored English in the zero-shot setting (accuracy 92% vs 76%, p < 0.01). For robust features such as ECOG PS, histology, and brain metastases, both languages achieved comparably high accuracy and F1-scores, with no statistically significant differences (p > 0.05). Across all prompting strategies and features, performance on T, N, and Stage remained low, regardless of language, though Italian showed marginal improvements (e.g., Stage F1-score 0.38 vs 0.43 in zero-shot, p < 0.01).

**Figure 3:**
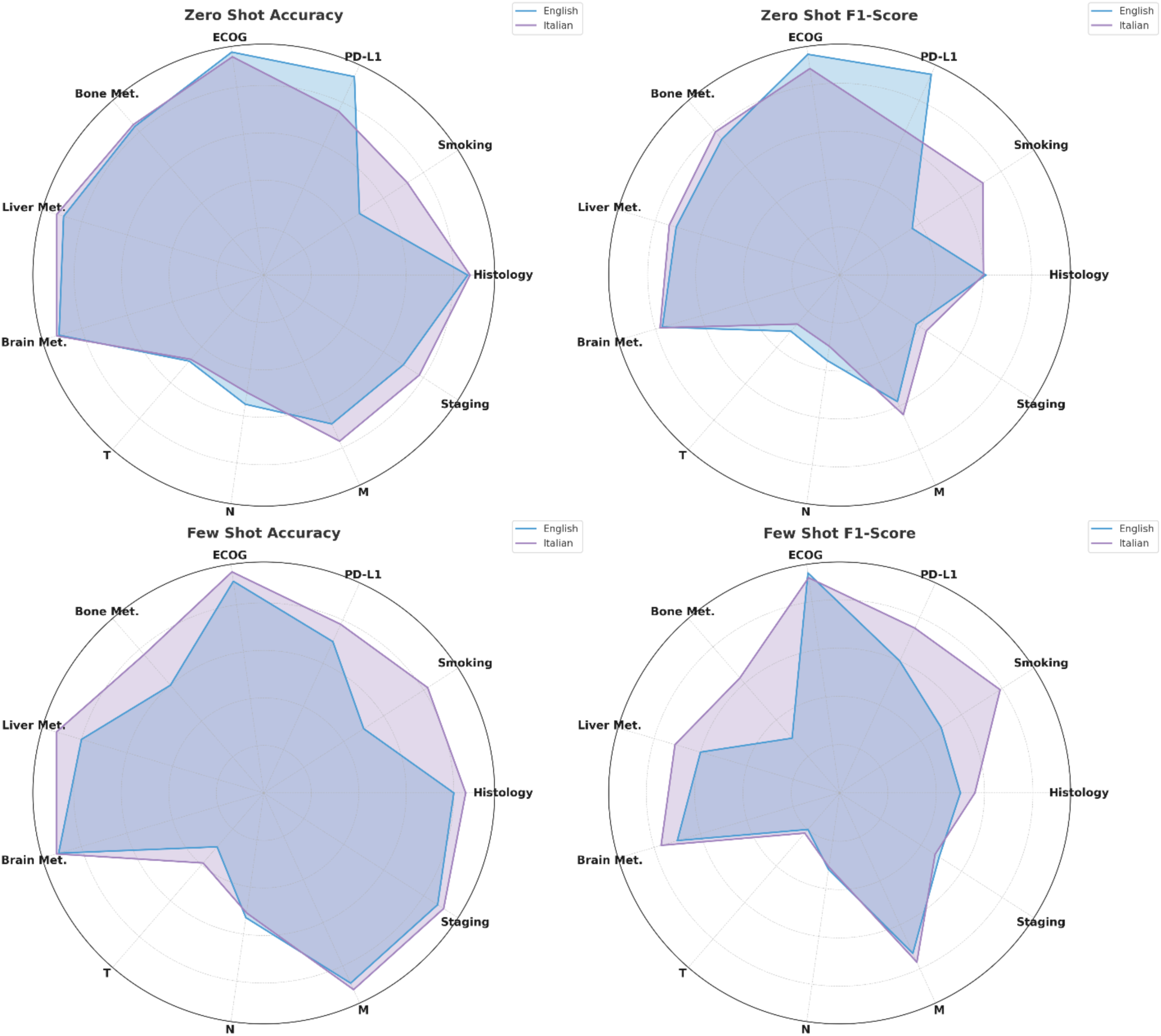
Spider plots of LLaMa 3.1 performance in a) accuracy and b) F1-score across all features extracted with English and Italian prompts.

#### 3.2.3 Agreement analysis between models

To systematically quantify agreement between different LLMs and prompting strategies, we computed pairwise Fleiss’ Kappa scores across all cancer-related features. As shown in Figure S5.2 the distributions of κ values vary notably across model-prompt combinations. LLaMa 3.1 demonstrated the highest overall agreement when prompted with few-shot examples, with median κ values approaching the agreement threshold (κ ≈ 0.6–0.7). In contrast, Mistral models, particularly under zero-shot settings, exhibited both lower median κ and higher variability, including outliers indicating disagreement scores (κ < 0). Several comparisons reveal statistically significant differences after FDR correction (Tables S5.1 and S5.2). For instance, when using LLaMa 3,1 zero-shot as reference, agreement was significantly lower with Mistral zero (p = 0.0068) and Mistral few-shot with annotations (p = 0.00098), confirming that model and prompting strategy both substantially affect alignment. Notably, LLaMa few-shot prompting yielded significantly improved agreement over Mistral few-shot (*p* = 0.0049) and over its own baseline (*p* = 0.0049), confirming the additive benefit of in-context examples. Interestingly, adding annotation to Mistral few-shot did not uniformly increase agreement: although the median κ slightly improved, the variability remained high, and the statistical benefit was less pronounced (*p* = 0.0198 vs. Mistral few). These results indicate that LLaMa 3.1 not only achieves higher performance in isolation but also aligns more closely across prompting strategies, especially when guided by few-shot examples and structured annotations.

#### 3.2.5 Clinical experiment results

To investigate the influence of clinical expertise on prompt design and model output agreement, we compared Fleiss’ Kappa values (Tables S7.1 and S7.2) across three rater groups (i.e., experienced clinicians, residents, and medical students) under both zero-shot and few-shot prompting conditions (Figure 4). Our findings revealed that expertise level significantly impacted the consistency of information extracted using LLMs. Specifically, experienced clinicians showed higher and more stable agreement, while students exhibited greater variability. Notably, few-shot prompting led to statistically significant changes (Table S7.3) in agreement within the student group (Wilcoxon p-value = 0.031), and in the inter-group comparison between experienced and resident raters (Wilcoxon p-value = 0.047). Specifically, few-shot prompting slightly reduced agreement among experienced raters, while it appeared to improve consistency for residents and students. This suggests that few-shot prompting may provide beneficial scaffolding for less experienced users, potentially by reducing ambiguity in task interpretation. No significant changes were observed within experienced and resident groups or in other inter-group comparisons.

**Figure 4:**
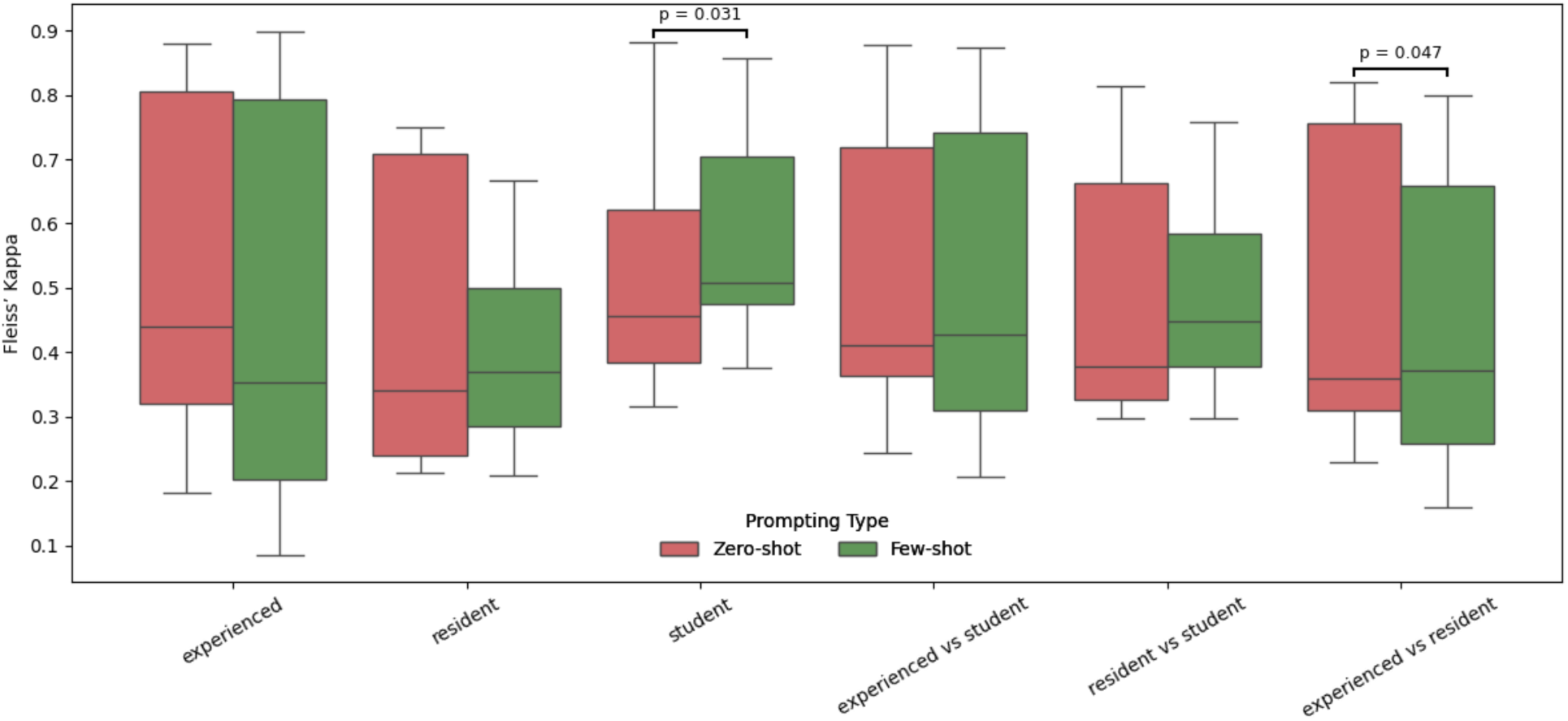
Agreement among clinical raters stratified by level of expertise and prompting strategy. Fleiss’ Kappa values are compared across six rater groups (intra- and inter-group) under zero-shot and few-shot prompting conditions. Statistically significant differences (Wilcoxon test, *p* < 0.05) are annotated above group pairs.

### 3.3 TKI cohort results

Heatmaps in Figure S8.1 compare the performance on the TKI cohort of LLaMa 3.1 and Mistral for all the cancer entities extracted. The few-shot+ann data strategy consistently yielded the highest performance across most clinical features for both LLaMa 3.1 8B and Mistral 7B. For LLaMa 3.1, this configuration produced higher scores in histology 0.99 (0.97-1.00), PD-L1 0.68 (0.61-0.85), ECOG PS 0.95 (0.92-0.98), and brain metastases 0.85 (0.80-0.90) (Table S3.3). The few-shot + annotation strategy improved Mistral’s performance in several features, notably T 0.32 (0.25-0.39), N 0.44 (0.37-0.52), and M 0.44 (0.39-0.53) descriptors, outperforming both its zero-shot [T: 0.24 (0.18-0.30), N: 0.20 (0.13-0.27), M: 0.28 (0.22-0.35) and few-shot-only counterparts (Table S3.4). For LLaMa 3.1, few-shot and few shot+ann. strategies produce markedly higher F1-scores for both categorical and binary variables. In smoking status, the F1-score reaches 0.82 (0.76-0.88) with few-shot, 0.78 (0.71-0.84) with few-shot+ann, but drops significantly to 0.51 (0.42-0.59) in the zero-shot configuration. Similarly, PD-L1 expression achieves an F1-score of 0.66 (0.58-0.73) with few shot+ann. data, while zero-shot decreases to 0.57 (0.52-0.62). Mistral shows slightly smaller gains from example-based prompting. In the TNM staging classification features, the F1-score always improves from zero-shot to few-shot+ann.

## 4. Discussion

Our work provides a comprehensive evaluation of how LLMs are used to structure data from EHRs of cancer patients. We benchmarked four privacy-preserving SLMs on the real-world dataset APOLLO 11^35^ of 711 NSCLC patients treated with IO and validated on a dataset of 186 NSCLC patients treated with TKIs. LLaMA 3.1 8B emerged as the best model with an average accuracy of 0.81 and F1-score of 0.62, substantially surpassing Mistral 7B (accuracy: 0.69, F1: 0.54), BioMistral 7B (accuracy: 0.44, F1: 0.29), and MedLLaMA 2 7B (accuracy: 0.27, F1: 0.24). Several factors may explain this counterintuitive finding. General-purpose LLMs, trained on diverse corpora, balance medical knowledge with general reasoning. Biomedical LLMs, though fine-tuned on benchmarks, risk overfitting to narrow domains. Alber et al. (2025)^36^ showed GPT-4 outperformed biomedical models on MedAlign, while Dorfner et al. (2025)^37^ found BioMedLM and Med-PaLM 2 underperformed in multi-step or cross-domain reasoning. Data mismatches further limit domain models, which struggle on unseen tasks, whereas generalist models remain robust. Indeed, many biomedical LLMs are tuned on public benchmarks that may overlap with generalist training data, yet these benchmarks fail to reflect real clinical complexity. As a result, domain models often struggle on unseen data, while generalist models remain robust. SLMs performance in extracting clinical variables varied notably by variable nature and context. Multiclass variables, which made up most of the study, were harder to capture than binary ones. Differences across cancer entities suggested that explicitly stated features were recalled more reliably than implied or context-dependent ones, indicating limits in contextual and implicit knowledge capture. Prompting experiments further showed that design strongly influenced outcomes, with example-driven approaches improving both accuracy and F1-scores in complex EHR texts. Indeed, the good performance obtained with sites of metastases in zero-shot settings suggests that SLMs can effectively infer true/false labels effectively. In contrast, for multiclass variables like histology, ECOG PS, PD-L1 expression and smoking there is the need for tailored prompting strategies. Extracting the T, N, M staging component remains a major challenge for SLMs. Our EHRs use complex cancer terminology, with staging rarely expressed explicitly (e.g., “T2”, “N1”) but instead through descriptive phrases such as “6 cm Squamous Intraepithelial Lesion” or “thoracic wall involvement.” Similarly, nodal and metastatic status are described indirectly (e.g., “32×19 mm subcarinal adenopathy,” “right adrenal lesion”), requiring mapping to categories using clinical judgment and guideline knowledge (RECIST^38^, AJCC^39^). Even with advanced prompting, both general-purpose and biomedical LLMs showed consistently low performance in this task. A subanalysis on pathology reports describing pT and pN has been conducted, demonstrating almost perfect extraction performance when this information is retrieved from structured reports instead of free-texts such as EHRs. These findings highlight the gap between free-text expression and structured collection of TNM staging, underscoring the need for human supervision or hybrid systems combining language models with rule-based validation pipelines. Chang et al. (2024)^40^ showed that open-source LLMs such as Med42-70B and ClinicalCamel-70B achieve modest performance (F1 score of 0.50) in T, N, M staging extraction even on well-curated datasets.. Our results found that LLMs benefit from annotated few-shot prompts also in challenging tasks such as T, M, and overall staging. For instance, BioMistral 7B improved its staging accuracy from 0.32 (zero-shot) to 0.59 with annotated prompting, while the same setting improved M extraction from 0.12 to 0.35. F1-score analysis further confirms that few-shot prompting with annotated data improves structured information extraction in clinical narratives. Huang et al. (2024)^41^ found that ChatGPT often misclassified TNM stages due to guideline misinterpretation and ambiguous language, though few-shot prompts with annotated examples improved accuracy. Similarly, Fung et al. (2025)^42^ showed that embedding structured examples from labeled datasets into prompts significantly boosted performance, especially for contextually complex tasks. Our comparison between English and Italian prompts revealed that native-language few-shot prompting leads to improved performance, particularly for complex or implicitly expressed clinical variables. Few-shot prompting with Italian examples significantly boosted performance on tasks such as staging, PD-L1, T, N, and M. Indeed, these variables often require interpretation of nuanced clinical language that is best captured in the original reporting language. Grothey et al. (2024),^43^ demonstrated that multilingual LLM performance declines markedly in non-English settings unless language-specific annotations or native-language prompts are used. In addition, we observed that inter-model agreement varied substantially across architectures and prompting strategies. LLaMa 3.1 with few-shot prompting consistently achieved the highest Fleiss κ scores, nearing the threshold for substantial agreement (κ ≈ 0.6–0.7), while Mistral models, especially in zero-shot settings, exhibited lower agreement and higher variability. Few-shot prompting notably enhanced alignment within and across models, with statistically significant gains observed for LLaMa 3.1 over Mistral. This is in line with findings by Mukherjee et al. (2023)^44^, who reported improved agreement using structured prompts with Vicuna-13B. In accordance with the prompting analysis, few-shot+ann reduced considerably the hallucination rate in case of missing features. As an extra analysis (Table S6.1 and Table S6.2), we labeled 8 of 11 features as *“not mentioned”* to ensure the LLM inferred MI only when no evidence appeared in the text. Unlike general web data where *“not mentioned”* implies absence, cancer entity recognition requires inference beyond explicit statements.

A novelty in our work is represented by the experiment on clinical expertise. In this analysis, we involved 26 medical participants, including experienced oncologists, resident doctors and medical students. To the best of our knowledge, this is the first study to investigate how the level of clinical experience affects prompt design and the consistency of LLM-assisted extraction. Our results reveal that expertise level strongly influences annotation agreement. Experienced clinicians showed higher and more stable agreement across both prompting strategies, whereas students exhibited greater variability. Notably, few-shot prompting significantly improved agreement among students, suggesting that worked examples may provide help to less experienced raters. These findings suggest that prompt design strategies should be tailored not only to the model, but also to the annotator’s expertise, particularly in human-in-the-loop pipelines for clinical information extraction. Finally, we validated our benchmarking pipeline on the TKI cohort. Only LLaMA 3.1 and Mistral have been tested in this analysis. The results obtained were consistent emphasizing the generalizability of our approach. From a clinical point of view, this study confirms that with minimal annotation effort, an LLM pipeline can be quickly adapted to diverse patient populations, making it a promising tool for scalable cancer data extraction in research and clinical practice. Overall, our study is generalizable to other cancer studies, since clinical features here extracted are common to many cancer types. Moreover, our pipeline, built on lightweight language models and local deployment, not only guarantees patient privacy, but also runs efficiently on modest computing infrastructures. In practical applications, SLMs provide several advantages, especially when integrated with preprocessing techniques, such as regular expressions, to highlight critical data in clinical notes.^45^ Furthermore, the use of SLM significantly reduces the computational burden associated with larger models, making them more suitable for healthcare settings where resources are limited. In our analysis, LLaMA 3.1 8B showed the strongest information retrieval while securing sensitive data. Models like GPT, with larger pre-training, can perform better but GPT-4’s cloud deployment limits use in personal healthcare.^46^ Open-source models, by contrast, allow freezing, auditing, and replicating behavior, unlike opaque proprietary systems.^20^ Fairness in LLM outputs is a major ethical concern, as models trained on non-diverse data may perpetuate healthcare disparities, leading to misdiagnosis or inappropriate treatment.^46^ Freyer et al. (2024)^48^ note a regulatory gap: while frameworks like the European AI Act^49^ exist, they are often poorly implemented. Tailored regulation is needed to address LLM-specific risks (e.g., hallucinations, variability, data provenance) and to distinguish general-purpose from health-specific LLMs, the latter falling under medical device rules.^51^ More broadly, foundation models in healthcare demand new evaluation methods to ensure safety, robustness, and clinical trustworthiness.^50^

Our analysis potentially helps clinicians in decision support systems, aiding in identifying cancer conditions. Enhancing information extraction from free text not only improves quantitative analysis in research, but it can streamline quality control in data recruitment, thereby reducing labor-intensive information extraction tasks. A recent study^52^ has proven LLM-assisted methods significantly reduced processing time for data extraction (from 86.9 minutes to 14.7 minutes per RCT) and could help assess the methodological quality of RCTs. This study also emphasizes the interaction between domain expertise and SLMs. Our experiments involving clinicians reflect not only the role of expertise on the SLM performance but also suggest that differences in performance may be partly influenced by clinicians’ attitudes toward AI and their familiarity in using these tools effectively. Moreover, clinicians should help in shaping how LLMs are used, ensuring that systems are not only accurate but also clinically acceptable and usable. This approach is the key to build trust and ensure clinical usability.^53^ Overall, aligning medical documentation with LLM best practices can enhance both clinical communication and automated data extraction. Ongoing research and the development of more sophisticated methods are essential to better understand and improve how these models operate. Nonetheless, this study has limitations. Results on features such as TNM staging need more investigation. Further refinement through fine-tuning or Retrieval Augmented Generation approaches could be necessary to overcome the tendency of SLMs to hallucinate.^54^ Our proof-of-concept study on clinical expertise included only a small group of oncologists, therefore future investigations will involve a larger cohort of healthcare professionals to confirm our results. Similarly, we plan to extend the set of clinical features including medications, timelines, side effects and comorbidities. In conclusion, our study provides a comprehensive test of SLMs for extensive extraction of structured information from EHRs that can be used for future investigations of SLMs in the cancer domain.

## Data Availability

All data produced in the present study are not readily available because of patients' privacy protection. Requests to access the datasets should be directed to the corresponding author

## Authors Contributions

F.C., V.P. contributed to concept/writing/data analysis; L.M. contributed to concept/writing; L.Passos., G.L., A.F. contributed to data curation; V.M. contributed to revision; A.P., P.B., A.L.G.P., F.d.B. contributed to concept/revision. J.A. contributed to data anonymization. I.C.W., F.W., J.N.K, contributed to concept. G.M., R.R., P.A., T.C., S.N., S.R., P.A., M.D.P., A.R., G.S., L.Provenzano, A.S., M.M.P., C.C., C.G., R.S., G.D.L., M.O., M.Brambilla, T.B., G.C., T.B., C.P., G.L.R. contributed to user study. M.Borracino, C.B., R.M.D.M., C.A., A.D.D., M.G. contributed to data collection.

## Declaration of Competing Interest

LM declares conference grants from Sanofi, Daiichi Sankyo, LEOPharma; honoraria from Novartis, MSD, Elma Research. VM declares speaker honorarium from Novartis. AF declares speaker honorarium from Novartis. L.Provenzano declares speaker honorarium from Novartis, Pfizer, Gilead. AS declares speaker honorarium from Novartis, MSD, BMS, Immunocore, Pierre Fabre. GC declares speaker honorarium from Takeda, conference grants from Amgen, AstraZeneca. TB declares conference grants from MSD, Sanofi, Pfizer, Lilly; honoraria from MSD. MO declares consulting/advisory role for AstraZeneca, BMS, MSD, Pfizer, J&J; honoraria from AstraZeneca, BMS, MSD; conference grants from Eli Lilly, J&J. M.Brambilla declares conference grants from Eli Lilly, J&J, LeoPharma, honoraria from BMS, AstraZeneca. CP declares personal fees from Italfarmaco, AstraZeneca, BMS, Merck Sharp and Dohme, and Janssen; and institutional funding from Novartis. JNK declares consulting services for Panakeia, AstraZeneca, MultiplexDx, Mindpeak, Owkin, DoMore Diagnostics, and Bioptimus. Furthermore, he holds shares in StratifAI, Synagen, Tremont AI, and Ignition Labs, has received an institutional research grant from GSK, and has received honoraria from AstraZeneca, Bayer, Daiichi Sankyo, Eisai, Janssen, Merck, MSD, BMS, Roche, Pfizer, and Fresenius. ALGP declares co-founder and shareholder of two startup companies, Agade srl and AllyArm srl; speaker honorarium from Novartis. FdB declares a patent for PCT/ IB2020/055956 pending and a patent for IT201900009954 pending; honoraria from, or consultant role for, Roche, EMD Serono, NMS Nerviano Medical Science, Sanofi, MSD, Novartis, Incyte, BMS, Menarini Healthcare Research & Pharmacoepidemiology, Merck Group, Pfizer, Servier, AMGEN, Incyte, outside the submitted work. GLR declares consultant role for Roche, Novartis, BMS, MSD, AstraZeneca, Takeda, Amgen, Sanofi, Italfarmaco, Pfizer; payment or honoraria for lectures, presentations, speakers bureaus, manuscript writing or educational events from Roche, Novartis, BMS, MSD, AstraZeneca, Takeda, Amgen, Sanofi; support for attending meetings and/or travel from Roche, BMS, MSD; data safety monitoring board or advisory board for Roche, Novartis, BMS, MSD, AstraZeneca, Sanofi; has acted as principal investigator in sponsored clinical trials for Roche, Novartis, BMS, MSD, AstraZeneca, GSK, Amgen, Sanofi, outside the submitted work. AP declares consulting/advisory role for BMS, AstraZeneca, Novartis; travel, accommodations, or other expenses paid or reimbursed by Roche, Italfarmaco; principal investigator of Spectrum Pharmaceuticals; personal fees from Roche, AstraZeneca and BMS, outside the submitted work. FC, VP, GL, L.Passos, JA, ICW, FW, GM, RR, PA, TC, SN, SR, PA, MDP, AR, GS, MMP, CC, CG, RS, M.Borracino., CB, RMDM, CA, ADD, GDL, MG, PB declare they have no financial or non-financial interests to disclose.

## Acknowledgments

We thank all the patients and the participating centers who accepted to actively participate in the Apollo 11 study with protocol number INT 128/22. This work was supported by Fondazione IRCCS ‘Istituto Nazionale dei Tumori’, 5 per 1000 Ministry of Health funds under the project ‘DWH 2.0: maintenance, integration, harmonization, and diffusion’; and 5 per 1000 Ministry of University and Research funds and Fondazione IRCSS Istituto Nazionale dei Tumori, through its call for the Valorisation of Institutional Research Program BRI 2021.

JNK is supported by the German Cancer Aid DKH (DECADE, 70115166), the German Federal Ministry of Research, Technology and Space BMFTR (PEARL, 01KD2104C; CAMINO, 01EO2101; TRANSFORM LIVER, 031L0312A; TANGERINE, 01KT2302 through ERA-NET Transcan; Come2Data, 16DKZ2044A; DEEP-HCC, 031L0315A; DECIPHER-M, 01KD2420A; NextBIG, 01ZU2402A), the German Research Foundation DFG (CRC/TR 412, 535081457; SFB 1709/1 2025, 533056198), the German Academic Exchange Service DAAD (SECAI, 57616814), the German Federal Joint Committee G-BA (TransplantKI, 01VSF21048), the European Union EU’s Horizon Europe research and innovation programme (ODELIA, 101057091; GENIAL, 101096312), the European Research Council ERC (NADIR, 101114631), the National Institutes of Health NIH (EPICO, R01 CA263318) and the National Institute for Health and Care Research NIHR (Leeds Biomedical Research Centre, NIHR203331). The views expressed are those of the author(s) and not necessarily those of the NHS, the NIHR or the Department of Health and Social Care. This work was funded by the European Union. Views and opinions expressed are however those of the author(s) only and do not necessarily reflect those of the European Union. Neither the European Union nor the granting authority can be held responsible for them.

## Data availability

The datasets presented in this article are not readily available because of patients’ privacy protection. Requests to access the datasets should be directed to the corresponding author.

## Notes

### Competing Interest Statement

All authors have completed the ICMJE uniform disclosure form at www.icmje.org/coi_disclosure.pdf and declare: no support from any organisation for the submitted work; LM declares conference grants from Sanofi, Daiichi Sankyo, LEOPharma; honoraria from Novartis, MSD, Elma Research. VM declares speaker honorarium from Novartis. AF declares speaker honorarium from Novartis. L.Provenzano declares speaker honorarium from Novartis, Pfizer, Gilead. AS declares speaker honorarium from Novartis, MSD, BMS, Immunocore, Pierre Fabre. GC declares speaker honorarium from Takeda, conference grants from Amgen, AstraZeneca. TB declares conference grants from MSD, Sanofi, Pfizer, Lilly; honoraria from MSD. MO declares consulting/advisory role for AstraZeneca, BMS, MSD, Pfizer, JeJ; honoraria from AstraZeneca, BMS, MSD; conference grants from Eli Lilly, JeJ. M.Brambilla declares conference grants from Eli Lilly, JeJ, LeoPharma, honoraria from BMS, AstraZeneca. CP declares personal fees from Italfarmaco, AstraZeneca, BMS, Merck Sharp and Dohme, and Janssen; and institutional funding from Novartis. JNK declares consulting services for Panakeia, AstraZeneca, MultiplexDx, Mindpeak, Owkin, DoMore Diagnostics, and Bioptimus. Furthermore, he holds shares in StratifAI, Synagen, Tremont AI, and Ignition Labs, has received an institutional research grant from GSK, and has received honoraria from AstraZeneca, Bayer, Daiichi Sankyo, Eisai, Janssen, Merck, MSD, BMS, Roche, Pfizer, and Fresenius. ALGP declares co-founder and shareholder of two startup companies, Agade srl and AllyArm srl; speaker honorarium from Novartis. FdB declares a patent for PCT/ IB2020/055956 pending and a patent for IT201900009954 pending; honoraria from, or consultant role for, Roche, EMD Serono, NMS Nerviano Medical Science, Sanofi, MSD, Novartis, Incyte, BMS, Menarini Healthcare Research & Pharmacoepidemiology, Merck Group, Pfizer, Servier, AMGEN, Incyte, outside the submitted work. GLR declares consultant role for Roche, Novartis, BMS, MSD, AstraZeneca, Takeda, Amgen, Sanofi, Italfarmaco, Pfizer; payment or honoraria for lectures, presenta- tions, speakers bureaus, manuscript writing or educational events from Roche, Novartis, BMS, MSD, AstraZeneca, Takeda, Amgen, Sanofi; support for attending meetings and/or travel from Roche, BMS, MSD; data safety monitoring board or advisory board for Roche, Novartis, BMS, MSD, AstraZeneca, Sanofi; has acted as principal investigator in spon- sored clinical trials for Roche, Novartis, BMS, MSD, AstraZeneca, GSK, Amgen, Sanofi, outside the submitted work. AP declares consulting/advisory role for BMS, AstraZeneca, Novartis; travel, accommodations, or other expenses paid or reimbursed by Roche, Italfarmaco; principal investigator of Spectrum Pharmaceuticals; personal fees from Roche, AstraZeneca and BMS, outside the submitted work.
FC, VP, GL, L.Passos, JA, ICW, FW, GM, RR, PA, TC, SN, SR, PA, MDP, AR, GS, MMP, CC, CG, RS, M.Borracino., CB, RMDM, CA, ADD, GDL, MG, PB declare they have no financial or non-financial interests to disclose.
No other relationships or activities that could appear to have influenced the submitted work.

### Clinical Trial

NCT05550961

### Clinical Protocols

https://clinicaltrials.gov/study/NCT05550961

### Funding Statement

This study was funded by Fondazione IRCCS Istituto Nazionale dei Tumori di Milano, 5 per 1000 Ministry of Health funds under the project DWH 2.0: maintenance, integration, harmonization, and diffusion; and 5 per 1000 Ministry of University and Research funds and Fondazione IRCSS Istituto Nazionale dei Tumori, through its call for the Valorisation of Institutional Research Program BRI 2021.

### Author Declarations

Ethics committee/IRB of Fondazione IRCCS Istituto Nazionale dei Tumori di Milano gave ethical approval for this work

