## Supplementary material for "Combining Clinician Expertise with Prompt Engineering enhances Small Language Models Reliability for Cancer Entity Recognition in Electronic Health Records"

#### S1 Study cohorts

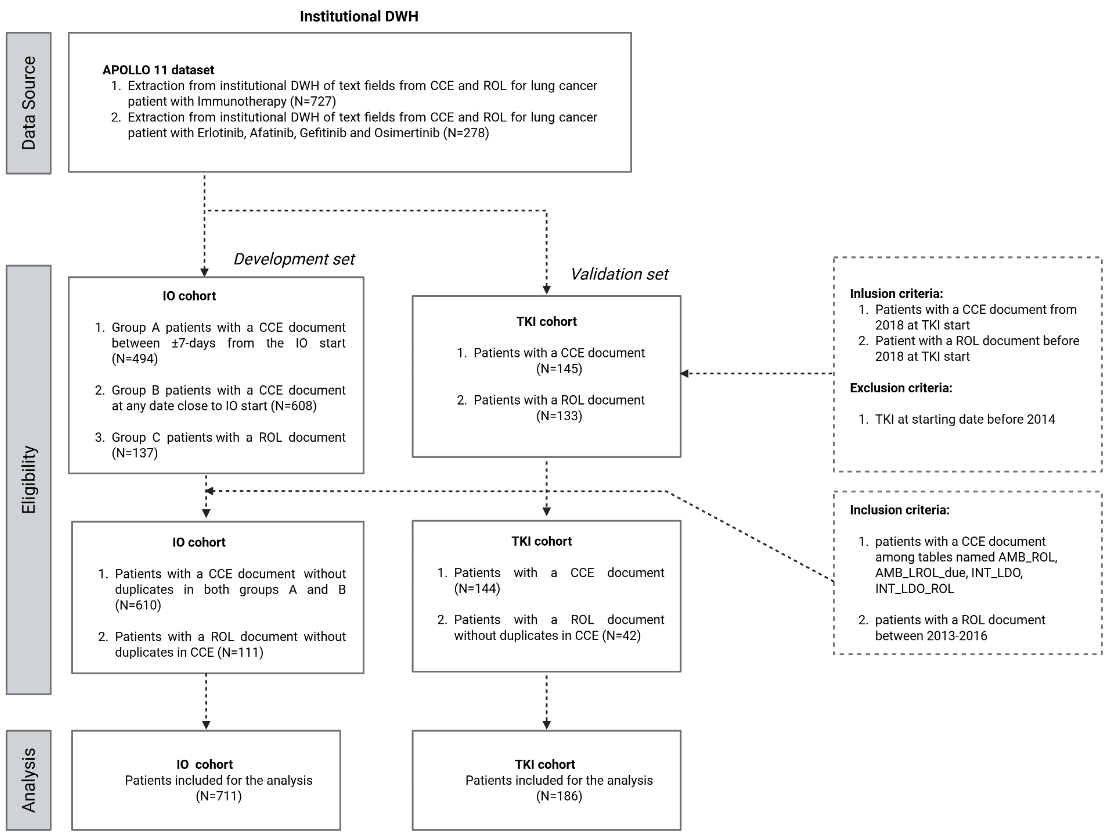


**Figure S1.1:** Flowchart of the patient selection from the institutional DWH. Data are split into development (IO cohort) and validation set (TKI cohort).

Grammar: Example of JSON Output

```
{ "SMOKING": "former smoker" | "current smoker" | "never smoked" | "not mentioned",
"PDL1": "<1%" | "1-49%" | ">=50%" | "not mentioned",
"HISTOLOGY": "adenocarcinoma" | "squamous" | "other histology" | "not mentioned",
"ECOG": "0" | "1" | "2" | "3" | "not mentioned",
"T_IO_START": "T0" | "T1" | "T2" | "T3" | "T4" | "not mentioned",
"N_IO_START": "N0" | "N1" | "N2" | "N3" | "not mentioned",
"M_IO_START": "M0" | "M1",
"STAGE": "III" | "IV" | "not mentioned",
"BONE_METASTASIS": boolean,
"BRAIN_METASTASIS": boolean,
"LIVER_METASTASIS": boolean }
```

**Figure S2.1**: JSON grammar schema for all the features selected.

| Entity | Type | Labels |
|---|---|---|
| **Smoking history** | Multiclass | Current smoker, Former smoker, Never smoked, not mentioned |
| **Histological subtype** | Multiclass | Adenocarcinoma, Squamous cell carcinoma, Other histologies, Not mentioned |
| **ECOG PS** | Multiclass | 0 (Fully active), 1 (Restricted but ambulatory), 2 (Ambulatory, self-care possible), 3 (Limited selfcare, bed/chair-bound >50%), 4 (Completely disabled), 5 (Dead). |
| **PD-L1 expression** | Multiclass | <1%, 1–49%, >50%, Not mentioned |
| **Presence of Metastases** | Binary | Bone Metastases (Ture/False), Brain Metastases (True/False), Liver Metastases(True/False) |
| **Tumour staging (AJCC 8th Edition)** | Multiclass | T category (T0–T4), N category (N0–N3), M category (M0–M1), Not mentioned |
| **Overall clinical stage** | Multiclass | Stage III, Stage IV, not mentioned |

**Table S2.2**: Description of the cancer-related entities selected for the extraction.

### S3 Benchmark of models

**Table S3.1:** LlaMa 3.1 8B performance for each feature extracted and prompting strategy (Italian) on the IO cohort.

| Type of prompt | Metrics | Clinical Features | | | | | | | | | | |
|---|---|---|---|---|---|---|---|---|---|---|---|---|
| | | Histology | Smoking | PD-L1 | ECOG | Bone Met. | Liver Met. | Brain Met. | T | N | M | Stage |
| Zero-shot | Accuracy (%) | 0.87 [0.84, 0.89] | 0.72 [0.69, 0.75] | 0.76 [0.73, 0.79] | 0.93 [0.91, 0.95] | 0.84 [0.82, 0.87] | 0.91 [0.88, 0.93] | 0.91 [0.88, 0.93] | 0.47 [0.43, 0.50] | 0.50 [0.46, 0.54] | 0.77 [0.74, 0.80] | 0.78 [0.75, 0.81] |
| | F1 score | 0,60 [0.56, 0.65] | 0.71 [0.69, 0.77] | 0.66 [0.64, 0.69] | 0.87 [0.82, 0.92] | 0.79 [0.75, 0.83] | 0.74 [0.69, 0.80] | 0.78 [0.73, 0.83] | 0.27 [0.25, 0.37] | 0.30 [0.27, 0.35] | 0.64 [0.60, 0.68] | 0.43 [0.40, 0.46] |
| | Precision (%) | 0,73 [0.70, 0.77] | 0.80 [0.75, 0.83] | 0.85 [0.83, 0.87] | 0.86 [0.80, 0.92] | 0.85 [0.80, 0.89] | 0.70 [0.62, 0.78] | 0.74 [0.66, 0.80] | 0.43 [0.24, 0.46] | 0.40 [0.33, 0.47] | 0.63 [0.60, 0.66] | 0.42 [0.40, 0.44] |
| | Recall (%) | 0,78 [0.75, 0.80] | 0.68 [0.64, 0.72] | 0.71 [0.70, 0.73] | 0.88 [0.82, 0.93] | 0.74 [0.68, 0.79] | 0.79 [0.71, 0.86] | 0.84 [0.77, 0.89] | 0.29 [0.26, 0.32] | 0.31 [0.28, 0.34] | 0.82 [0.78, 0.86] | 0.56 [0.53, 0.58] |
| Few-shot | Accuracy (%) | 0.85 [0.82, 0.88] | 0.82 [0.79, 0.84] | 0.78 [0.75, 0.81] | 0.94 [0.93, 0.96] | 0.77 [0.74, 0.80] | 0.91 [0.89 , 0.93] | 0.91 [0.89, 0.93] | 0.39 [0.36, 0.43] | 0.51 [0.47, 0.54] | 0.91 [0.89, 0.93] | 0.90 [0.88, 0.92] |
| | F1 score | 0.56 [0.51, 0.61] | 0.79 [0.75, 0.82] | 0.75 [0.72, 0.78] | 0.90 [0.85, 0.94] | 0.63 [0.57, 0.68] | 0.71 [0.64, 0.78] | 0.77 [0.71, 0.82] | 0.22 [0.19, 0.25] | 0.31 [0.27, 0.34] | 0.77 [0.72, 0.82] | 0.47 [0.43, 0.50] |
| | Precision (%) | 0.72 [0.69, 0.75] | 0.77 [0.73, 0.81] | 0.84 [0.81, 0.87] | 0.90 [0.84, 0.95] | 0.91 [0.86, 0.96] | 0.81 [0.73, 0.89] | 0.82 [0.75, 0.88] | 0.24 [0.20, 0.27] | 0.40 [0.33, 0.46] | 0.75 [0.69, 0.80] | 0.47 [0.43, 0.52] |
| | Recall (%) | 0.76 [0.74, 0.77] | 0.82 [0.78, 0.86] | 0.75 [0.73, 0.78] | 0.90 [0.85, 0.94] | 0.48 [0.42, 0.54] | 0.63 [0.55, 0.72] | 0.72 [0.65, 0.79] | 0.24 [0.21, 0.27] | 0.31 [0.28, 0.34] | 0.81 [0.75, 0.86] | 0.46 [0.42, 0.50] |
| Few-shot + clinical ann. | Accuracy (%) | 0.91 [0.89, 0.93] | 0.87 [0.84, 0.89] | 0.83 [0.80, 0.85] | 0.97 [0.95, 0.98] | 0.82 [0.79, 0.85] | 0.91 [0.89, 0.93] | 0.90 [0.88, 0.93] | 0.48 [0.45, 0.52] | 0.43 [0.39, 0.47] | 0.92 [0.90, 0.94] | 0.91 [0.89, 0.93] |
| | F1 score | 0.75 [0.69, 0.80] | 0.85 [0.82, 0.88] | 0.81 [0.78, 0.84] | 0.95 [0.91, 0.97] | 0.76 [0.71, 0.80] | 0.73 [0.66, 0.79] | 0.76 [0.70, 0.81] | 0.20 [0.17, 0.23] | 0.26 [0.22, 0.30] | 0.73 [0.67, 0.79] | 0.41 [0.36, 0.45] |
| | Precision (%) | 0.72 [0.69, 0.76] | 0.88 [0.85, 0.91] | 0.86 [0.84, 0.89] | 0.94 [0.89, 0.98] | 0.85 [0.80, 0.89] | 0.76 [0.68, 0.83] | 0.77 [0.69, 0.83] | 0.27 [0.22, 0.32] | 0.42 [0.34, 0.49] | 0.79 [0.71, 0.85] | 0.52 [0.44, 0.59] |
| | Recall (%) | 0.90 [0.80, 0.95] | 0.83 [0.79, 0.87] | 0.81 [0.78, 0.83] | 0.95 [0.93, 0.97] | 0.68 [0.63, 0.74] | 0.71 [0.63, 0.79] | 0.74 [0.67, 0.81] | 0.22 [0.20, 0.24] | 0.28 [0.25, 0.31] | 0.70 [0.64, 0.75] | 0.39 [0.36, 0.42] |

**Table S3.2:** Mistral 7B performance for each feature extracted and prompting strategy (Italian) on the IO cohort.

| Type of prompt | Metrics | Clinical Features | | | | | | | | | | |
|---|---|---|---|---|---|---|---|---|---|---|---|---|
| | | Histology | Smoking | PD-L1 | ECOG | Bone Met. | Liver Met. | Brain Met. | T | N | M | Stage |
| Zero-shot | Accuracy (%) | 0.85 [0.82, 0.88] | 0.67 [0.64, 0.70] | 0.81 [0.78, 0.84] | 0.89 [0.86, 0.91] | 0.79 [0.76,0.82] | 0.80 [0.77, 0.83] | 0.86 [0.83, 0.88] | 0.24 [0.21, 0.27] | 0.47 [0.44, 0.51] | 0.68 [0.65, 0.71] | 0.79 [0.76, 0.82] |
| | F1 score | 0.54 [0.49, 0.59] | 0.57 [0.53, 0.62] | 0.78 [0.74, 0.80] | 0.87 [0.84, 0.90] | 0.67 [0.62, 0.72] | 0.58 [0.52, 0.64] | 0.70 [0.64, 0.75] | 0.19 [0.16, 0.22] | 0.35 [0.31, 0.39] | 0.55 [0.51, 0.59] | 0.42 [0.39, 0.45] |
| | Precision (%) | 0.72 [0.69, 0.75] | 0.64 [0.57, 0.71] | 0.84 [0.81, 0.87] | 0.85 [0.81, 0.89] | 0.89 [0.84, 0.93] | 0.46 [0.39, 0.52] | 0.60 [0.53, 0.67] | 0.33 [0.25, 0.40] | 0.36 [0.33, 0.40] | 0.57 [0.55, 0.60] | 0.41 [0.39, 0.43] |
| | Recall (%) | 0.75 [0.73, 0.76] | 0.58 [0.55, 0.63] | 0.79 [0.76, 0.81] | 0.91 [0.88, 0.93] | 0.54 [0.48, 0.60] | 0.80 [0.72, 0.87] | 0.84 [0.77, 0.90] | 0.27 [0.21, 0.36] | 0.41 [0.33, 0.54] | 0.69 [0.63, 0.75] | 0.49 [0.44, 0.53] |
| Few-shot | Accuracy (%) | 0.77 [0.74, 0.80] | 0.65 [0.62, 0.68] | 0.89 [0.87, 0.92] | 0.87 [0.84, 0.89] | 0.77 [0.74, 0.81] | 0.87 [0.84, 0.89] | 0.85 [0.83, 0.88] | 0.23 [0.20, 0.25] | 0.46 [0.42, 0.50] | 0.65 [0.62, 0.69] | 0.85 [0.82, 0.87] |
| | F1 score | 0.49 [0.47, 0.51] | 0.56 [0.51, 0.61] | 0.89 [0.87, 0.92] | 0.86 (95% CI: [0.82, 0.89]) | 0.70 [0.66, 0.75] | 0.67 [0.60, 0.73] | 0.69 [0.63, 0.74] | 0.16 [0.14, 0.18] | 0.33 [0.30, 0.36] | 0.54 [0.50, 0.57] | 0.47 [0.42, 0.52] |
| | Precision (%) | 0.72 [0.47, 0.74] | 0.71 [0.62, 0.77] | 0.90 [0.88, 0.92] | 0.86 [0.84, 0.88] | 0.74 [0.69, 0.80] | 0.59 [0.52, 0.67] | 0.60 [0.53, 0.67] | 0.24 [0.21, 0.27] | 0.34 [0.30, 0.36] | 0.58 [0.55, 0.60] | 0.46 [0.42, 0.50] |
| | Recall (%) | 0.71 [0.69, 0.72] | 0.59 [0.55, 0.62] | 0.89 [0.87, 0.91] | 0.89 [0.83, 0.92] | 0.67 [0.61, 0.72] | 0.77 [0.70, 0.85] | 0.81 [0.74, 0.87] | 0.23 [0.20, 0.26] | 0.36 [0.32, 0.39] | 0.71 [0.66, 0.77] | 0.52 [0.42, 0.68] |
| Few-shot + clinical ann. | Accuracy (%) | 0.88 [0.86, 0.90] | 0.64 [0.60, 0.68] | 0.83 [0.81, 0.86] | 0.90 [0.88, 0.93] | 0.45 [0.41, 0.49] | 0.85 [0.82, 0.87] | 0.87 [0.85, 0.90] | 0.21 [0.18, 0.24] | 0.48 [0.45, 0.52] | 0.73 [0.69, 0.76] | 0.84 [0.81, 0.86] |
| | F1 score | 0.68 [0.63, 0.73] | 0.59 [0.53, 0.63] | 0.83 [0.81, 0.86] | 0.88 [0.84, 0.91] | 0.59 [0.55, 0.62] | 0.63 [0.57, 0.70] | 0.70 [0.64, 0.76] | 0.17 [0.14, 0.19] | 0.34 [0.30, 0.37] | 0.58 [0.54, 0.62] | 0.45 [0.40, 0.50] |
| | Precision (%) | 0.68 [0.64, 0.71] | 0.69 [0.62, 0.74] | 0.84 [0.81, 0.86] | 0.88 [0.86, 0.90] | 0.42 [0.38, 0.46] | 0.55 [0.47, 0.62] | 0.67 [0.59, 0.74] | 0.27 [0.24, 0.30] | 0.35 [0.32, 0.39] | 0.59 [0.56, 0.62] | 0.51 [0.46, 0.57] |
| | Recall (%) | 0.79 [0.65, 0.88] | 0.59 [0.55, 0.63] | 0.84 [0.81, 0.86] | 0.89 [0.84, 0.93] | 0.98 [0.96, 0.99] | 0.76 [0.68, 0.83] | 0.74 [0.66, 0.81] | 0.22 [0.19, 0.25] | 0.36 [0.32, 0.40] | 0.72 [0.66, 0.78] | 0.46 [0.37, 0.61] |

**Table S3.3:** LlaMa 3.1 8B performance for each feature extracted and prompting strategy (Italian) on the TKI cohort.

| Type of prompt | Metrics | Clinical Features | | | | | | | | | | |
|---|---|---|---|---|---|---|---|---|---|---|---|---|
| | | Histology | Smoking | PD-L1 | ECOG | Bone Met. | Liver Met. | Brain Met. | T | N | M | Stage |
| Zero-shot | Accuracy (%) | 0.98 [0.96, 1.00] | 0.53 [0.46, 0.60] | 0.64 [0.57, 0.70] | 0.92 [0.88, 0.96] | 0.89 [0.83, 0.92] | 0.88 [0.82, 0.92] | 0.83 [0.77, 0.88] | 0,11 [0.06, 0.16] | 0,21 [0.15, 0.27] | 0,48 [0.40, 0.55] | 0,27 [0.20, 0.33] |
| | F1 score | 0.83 [0.53, 1.00] | 0.51 [0.42, 0.59] | 0.57 [0.52, 0.62] | 0.9 [0.75, 0.96] | 0.89 [0.84, 0.93] | 0.6 [0.43, 0.74] | 0.77 [0.69, 0.85] | 0,1 [0.06, 0.14] | 0,22 [0.12, 0.34] | 0,26 [0.23, 0.30] | 0,19 [0.15, 0.24] |
| | Precision (%) | 0.94 [0.52, 1.00] | 0.7 [0.64, 0.77] | 0.54 [0.48, 0.59] | 0.88 [0.77, 0.96] | 0.86 [0.78, 0.92] | 0.46 [0.31, 0.62] | 0.66 [0.55, 0.76] | 0,07 [0.04, 0.10] | 0,14 [0.07, 0.26] | 0,23 [0.19, 0.26] | 0,46 [0.11, 0.49] |
| | Recall (%) | 0.78 [0.53, 1.00] | 0.51 [0.44, 0.58] | 0.62 [0.57, 0.68] | 0.93 [0.76, 0.96] | 0.93 [0.85, 0.96] | 0.85 [0.64, 0.95] | 0.93 [0.83, 0.97] | 0,25 [0.18, 0.32] | 0,47 [0.29, 0.63] | 0,44 [0.26, 0.61] | 0,5 [0.27, 0.64] |
| Few-shot | Accuracy (%) | 0.98 [0.96, 1.00] | 0.84 [0.78, 0.89] | 0.62 [0.55, 0.69] | 0.94 [0.90, 0.97] | 0.83 [0.77, 0.88] | 0.9 [0.85, 0.94] | 0.84 [0.78, 0.88] | 0,1 [0.06, 0.14] | 0,19 [0.14, 0.24] | 0,54 [0.47, 0.61] | 0,31 [0.24, 0.37] |
| | F1 score | 0.95 [0.83, 1.00] | 0.82 [0.76, 0.88] | 0.58 [0.50, 0.65] | 0.87 [0.76, 0.97] | 0.82 [0.75, 0.88] | 0.64 [0.45, 0.79] | 0.76 [0.68, 0.83] | 0,1 [0.05, 0.14] | 0,28 [0.12, 0.38] | 0,3 [0.23, 0.37] | 0,24 [0.16, 0.32] |
| | Precision (%) | 0.95 [0.81, 1.00] | 0.86 [0.79, 0.91] | 0.8 [0.69, 0.87] | 0.87 [0.76, 0.97] | 0.92 [0.84, 0.96] | 0.53 [0.36, 0.70] | 0.7 [0.58, 0.79] | 0,07 [0.03, 0.11] | 0,19 [0.07, 0.33] | 0,23 [0.19, 0.29] | 0,48 [0.11, 0.54] |
| | Recall (%) | 0.95 [0.82, 1.00] | 0.8 [0.74, 0.87] | 0.57 [0.50, 0.64] | 0.87 [0.77, 0.98] | 0.73 [0.64, 0.81] | 0.8 [0.58, 0.92] | 0.84 [0.73, 0.91] | 0,24 [0.15, 0.33] | 0,57 [0.29, 0.64] | 0,48 [0.30, 0.65] | 0,55 [0.32, 0.67] |
| Few-shot + clinical ann. | Accuracy (%) | 0.99 [0.97, 1.00] | 0.78 [0.72, 0.83 | 0.68 [0.61, 0.75] | 0.95 [0.92, 0.98] | 0.89 [0.83, 0.92] | 0.86 [0.80, 0.90] | 0.85 [0.80, 0.90] | 0,1 [0.06, 0.15] | 0,2 [0.14, 0.26] | 0,57 [0.50, 0.64] | 0,32 [0.25, 0.39] |
| | F1 score | 0.62 [0.54, 1.00] | 0.78 [0.71, 0.84] | 0.66 [0.58, 0.73] | 0.92 [0.78, 0.98] | 0.89 [0.84, 0.93] | 0.55 [0.38, 0.69] | 0.79 [0.71, 0.87] | 0,06 [0.03, 0.09] | 0,24 [0.10, 0.36] | 0,25 [0.23, 0.26] | 0,17 [0.14, 0.26] |

| | | | | | | | | | | | | |
|---|---|---|---|---|---|---|---|---|---|---|---|---|
| | Precision (%) | 0.62 [0.52, 1.00] | 0.84 [0.79, 0.89] | 0.81 [0.73, 0.88] | 0.91 [0.79, 0.98] | 0.88 [0.80, 0.93] | 0.42 [0.28, 0.58] | 0.7 [0.59, 0.79] | 0,25 [0.02, 0.29] | 0,42 [0.08, 0.58] | 0,2 [0.17, 0.22] | 0,44 [0.10, 0.65] |
| | Recall (%) | 0.62 [0.54, 1.00] | 0.76 [0.69, 0.82] | 0.65 [0.58, 0.71] | 0.95 [0.77, 0.98] | 0.89 [0.83, 0.93] | 0.8 [0.58, 0.92] | 0.91 [0.81, 0.96] | 0,21 [0.20, 0.24] | 0,43 [0.27, 0.59] | 0,33 [0.32, 0.33] | 0,34 [0.33, 0.51] |

**Table S3.4:** Mistral 7B performance for each feature extracted and prompting strategy (Italian) on the TKI cohort.

| Type of prompt | Metrics | Clinical Features | | | | | | | | | | |
|---|---|---|---|---|---|---|---|---|---|---|---|---|
| | | Histology | Smoking | PD-L1 | ECOG | Bone Met. | Liver Met. | Brain Met. | T | N | M | Stage |
| Zero-shot | Accuracy (%) | 0.97 [0.94, 0.99] | 0.56 [0.48, 0.63] | 0.68 [0.61, 0.75] | 0.9 [0.85, 0.94] | 0.81 [0.75, 0.86] | 0.89 [0.84, 0.93] | 0.84 [0.79, 0.89] | 0,24 [0.18, 0.30] | 0,2 [0.13, 0.27] | 0,28 [0.22, 0.35] | 0,29 [0.23, 0.35] |
| | F1 score | 0.5 [0.33, 0.87] | 0.46 [0.38, 0.52] | 0.62 [0.56, 0.69] | 0.88 [0.74, 0.94] | 0.79 [0.48, 0.50] | 0.64 [0.48, 0.78] | 0.79 [0.71, 0.86] | 0,14 [0.09, 0.17] | 0,2 [0.13, 0.27] | 0,2 [0.16, 0.23] | 0,25 [0.19, 0.30] |
| | Precision (%) | 0.44 [0.33, 0.80] | 0.55 [0.38, 0.71] | 0.79 [0.74, 0.85] | 0.88 [0.76, 0.95] | 0.9 [0.46, 0.50] | 0.5 [0.34, 0.66] | 0.68 [0.46, 0.50] | 0,2 [0.17, 0.24] | 0,31 [0.22, 0.39] | 0,23 [0.19, 0.26] | 0,41 [0.33, 0.47] |
| | Recall (%) | 0.66 [0.32, 0.99] | 0.51 [0.44, 0.58] | 0.66 [0.59, 0.72] | 0.9 [0.73, 0.94] | 0.7 [0.49, 0.50] | 0.9 [0.70, 0.97] | 0.93 [0.49, 0.50] | 0,22 [0.16, 0.28] | 0,51 [0.21, 0.61] | 0,41 [0.15, 0.51] | 0,59 [0.26, 0.64] |
| Few-shot | Accuracy (%) | 0.99 [0.98, 1.00] | 0.55 [0.48, 0.62] | 0.74 [0.68, 0.80] | 0.74 [0.67, 0.80] | 0.83 [0.77, 0.88] | 0.91 [0.86, 0.95] | 0.87 [0.82, 0.91] | 0,16 [0.11, 0.21] | 0,22 [0.15, 0.28] | 0,36 [0.29, 0.44] | 0,42 [0.35, 0.49] |
| | F1 score | 0.56 [0.33, 1.00] | 0.45 [0.39, 0.52] | 0.71 [0.64, 0.77] | 0.64 [0.58, 0.79] | 0.82 [0.76, 0.88] | 0.68 [0.50, 0.81] | 0.81 [0.72, 0.87 | 0,1 [0.06, 0.14] | 0,22 [0.14, 0.30] | 0,24 [0.20, 0.27] | 0,37 [0.27, 0.46] |
| | Precision (%) | 0.5 [0.33, 1.00] | 0.56 [0.39, 0.72] | 0.82 [0.76, 0.88] | 0.73 [0.70, 0.89] | 0.86 [0.77, 0.92] | 0.57 [0.68, 0.87] | 0.75 [0.63, 0.84] | 0,21 [0.18, 0.24] | 0,33 [0.24, 0.40] | 0,24 [0.21, 0.27] | 0.47 [0.40, 0.54] |
| | Recall (%) | 0.67 [0.33, 1.00] | 0.52 [0.45, 0.59] | 0.69 [0.61, 0.76] | 0.61 [0.55, 0.76] | 0.79 [0.69, 0.86] | 0.85 [0.79, 0.96] | 0.88 [0.77, 0.94] | 0,22 [0.14, 0.34] | 0,54 [0.25, 0.62] | 0,53 [0.49, 0.56] | 0,71 [0.37, 0.73] |
| Few-shot + clinical ann. | Accuracy (%) | 0,98 [0.96, 1.00] | 0,47 [0.40, 0.54] | 0,74 [0.67, 0.81] | 0,76 [0.70, 0.82] | 0.67 [0.60, 0.74] | 0.88 [0.83, 0.92]) | 0.88 [0.82, 0.92] | 0,32 [0.25, 0.39] | 0,24 [0.18, 0.30] | 0,44 [0.37, 0.52] | 0,46 [0.39, 0.53] |
| | F1 score | 0,5 [0.33, 1.00] | 0,42 [0.35, 0.48] | 0,71 [0.62, 0.79] | 0,66 [0.60, 0.79] | 0.75 [0.57, 0.70] | 0.58 [0.41, 0.72] | 0.8 [0.72, 0.87] | 0,18 [0.13, 0.24] | 0,23 [0.16, 0.30] | 0,25 [0.21, 0.29] | 0,44 [0.29, 0.57] |

| | | | | | | | | | | | | |
|---|---|---|---|---|---|---|---|---|---|---|---|---|
| | Precision (%) | 0,44 [0.33, 1.00] | 0,55 [0.39, 0.66] | 0,79 [0.71, 0.87] | 0,75 [0.72, 0.90] | 0.61 [0.69, 0.81] | 0.47 [0.31, 0.64] | 0.78 [0.66, 0.87] | 0,24 [0.18, 0.33] | 0,39 [0.32, 0.45] | 0,22 [0.19, 0.25] | 0,5 [0.38, 0.63] |
| | Recall (%) | 0,66 [0.33, 1.00] | 0,5 [0.43, 0.56] | 0,67 [0.59, 0.75] | 0,62 [0.57, 0.76] | 0.96 [0.62, 0.72 | 0.75 [0.53, 0.89] | 0.82 [0.71, 0.90] | 0,28 [0.17, 0.40] | 0,54 [0.22, 0.61] | 0,42 [0.24, 0.59] | 0,61 [0.37, 0.75] |

### S4 Language experiment

**Table S4.1**: Performances of LlaMa 3.1 8B for each feature extracted and language of zero-shot prompting. **P*value of the McNemar test performed between the contingency tables for prompts in Italian (Ita) and in English (Eng).

| Metrics | Clinical Features | | | | | | | | | | | | | | | | | | | | | |
|---|---|---|---|---|---|---|---|---|---|---|---|---|---|---|---|---|---|---|---|---|---|---|
| | Histology | | Smoking | | PD-L1 | | ECOG | | Bone Met. | | Liver Met. | | Brain Met. | | T | | N | | M | | Stage | |
| | Eng | Ita | Eng | Ita | Eng | Ita | Eng | Ita | Eng | Ita | Eng | Ita | Eng | Ita | Eng | Ita | Eng | Ita | Eng | Ita | Eng | Ita |
| Accuracy (%) | 0,86 [0.84, 0.89] | 0.87 [0.84, 0.89] | 0.48 [0.45, 0.52] | 0.72 [0.69, 0.75] | 0,92 [0.90, 0.94] | 0.76 [0.73, 0.79] | 0,95 [0.93, 0.97] | 0.93 [0.91, 0.95] | 0.83 [0.80, 0.85] | 0.84 [0.82, 0.87] | 0,88 [0.86, 0.91] | 0.91 [0.88, 0.93] | 0.90 [0.8, 0.93] | 0.91 [0.88, 0.93] | 0.48 [0.45, 0.52] | 0.47 [0.43, 0.50] | 0.55 [0.51, 0.59] | 0.50 [0.46, 0.54] | 0.69 [0.66, 0.72] | 0.77 [0.74, 0.80] | 0.70 [0.67, 0.73] | 0.78 [0.75, 0.81] |
| F1 score | 0,61 [0.51, 0.70] | 0,60 [0.56, 0.65] | 0.36 [0.33, 0.40] | 0.71 [0.69, 0.77] | 0,92 [0.90, 0.94] | 0.66 [0.64, 0.69] | 0,93 [0.90, 0.96] | 0.87 [0.82, 0.92] | 0.75 [0.71, 0.79] | 0.79 [0.75, 0.83] | 0,71 [0.65, 0.76] | 0.74 [0.69, 0.80] | 0.77 [0.73, 0.82] | 0.78 [0.73, 0.83] | 0.31 [0.28, 0.35] | 0.27 [0.25, 0.37] | 0.36 [0.33, 0.42] | 0.30 [0.27, 0.35] | 0.58 [0.54, 0.62] | 0.64 [0.60, 0.68] | 0.38 [0.36, 0.41] | 0.43 [0.40, 0.46] |
| Precision (%) | 0,78 [0.67, 0.93] | 0,73 [0.70, 0.77] | 0.71 [0.46, 0.72] | 0.80 [0.75, 0.83] | 0,92 [0.90, 0.94] | 0.85 [0.83, 0.87] | 0,94 [0.88, 0.97] | 0.86 [0.80, 0.92] | 0.88 [0.84, 0.93] | 0.85 [0.80, 0.89] | 0,63 [0.56, 0.71] | 0.70 [0.62, 0.78] | 0.73 [0.66, 0.80] | 0.74 [0.66, 0.80] | 0.43 [0.30, 0.50] | 0.43 [0.24, 0.46] | 0.40 [0.36, 0.46] | 0.40 [0.33, 0.47] | 0.61 [0.58, 0.63] | 0.63 [0.60, 0.66] | 0.40 [0.38, 0.42] | 0.42 [0.40, 0.44] |
| Recall (%) | 0,6 [0.51, 0.73] | 0,78 [0.75, 0.80] | 0.52 [0.50, 0.55] | 0.68 [0.64, 0.72] | 0,91 [0.89, 0.93] | 0.71 [0.70, 0.73] | 0,94 [0.91, 0.96] | 0.88 [0.82, 0.93] | 0.65 [0.60, 0.71] | 0.74 [0.68, 0.79] | 0,80 [0.73, 0.87] | 0.79 [0.71, 0.86] | 0.83 [0.77, 0.89] | 0.84 [0.77, 0.89] | 0.32 [0.29, 0.35] | 0.29 [0.26, 0.32] | 0.36 [0.32, 0.41] | 0.31 [0.28, 0.34] | 0.80 [0.75, 0.83] | 0.82 [0.78, 0.86] | 0.52 [0.49, 0.55] | 0.56 [0.53, 0.58] |
| *p* value | <0.01 | | <0.01 | | <0.01 | | 0.98 | | 0.11 | | <0.05 | | 0,61 | | <0.01 | | <0.01 | | <0.01 | | <0.01 | |

**Table S4.2**: Performances of LlaMa 3.1 8B for each feature extracted and language of few-shot prompting. **P*value of the McNemar test performed between the contingency tables for prompts in Italian (Ita) and in English (Eng).

| Metrics | Clinical Features | | | | | | | | | | | | | | | | | | | | | |
|---|---|---|---|---|---|---|---|---|---|---|---|---|---|---|---|---|---|---|---|---|---|---|
| | Histology | | Smoking | | PD-L1 | | ECOG | | Bone Met. | | Liver Met. | | Brain Met. | | T | | N | | M | | Stage | |
| | Eng | Ita | Eng | Ita | Eng | Ita | Eng | Ita | Eng | Ita | Eng | Ita | Eng | Ita | Eng | Ita | Eng | Ita | Eng | Ita | Eng | Ita |
| Accuracy (%) | 0.86 [0.83, 0.88] | 0.85[0.82, 0.88] | 0.59 [0.56, 0.63] | 0.82 [0.79, 0.84] | 0.75 [0.71, 0.78] | 0.78 [0.75, 0.81] | 0.95 [0.93, 0.96] | 0.94 [0.93, 0.96] | 0,67 [0.63, 0.70] | 0.77 [0.74, 0.80] | 0,89 [0.87, 0.91] | 0.91 [0.89 , 0.93] | 0,90 [0.88, 0.92] | 0.91 [0.89, 0.93] | 0.35 [0.32, 0.39] | 0.39 [0.36, 0.43] | 0.53 [0.49, 0.56] | 0.51 [0.47, 0.54] | 0.88 [0.85, 0.90] | 0.91 [0.89, 0.93] | 0.87 [0.85, 0.90] | 0.90 [0.88, 0.92] |
| F1 score | 0.52 [0.47, 0.57] | 0.56 [0.51, 0.61] | 0.54 [0.50, 0.57] | 0.79 [0.75, 0.82] | 0.66 [0.64, 0.68] | 0.75 [0.72, 0.78] | 0.92 [0.88, 0.95] | 0.90 [0.85, 0.94] | 0,31 [0.25, 0.37] | 0.63 [0.57, 0.68] | 0,60 [0.53, 0.68] | 0.71 [0.64, 0.78] | 0,75 [0.69, 0.80] | 0.77 [0.71, 0.82] | 0.22 [0.19, 0.25] | 0.22 [0.19, 0.25] | 0.32 [0.29, 0.37] | 0.31 [0.27, 0.34] | 0.73 [0.68, 0.77] | 0.77 [0.72, 0.82] | 0.49 [0.46, 0.53] | 0.47 [0.43, 0.50] |
| Precision (%) | 0.47 [0.44, 0.51] | 0.72 [0.69, 0.75] | 0.69 [0.67, 0.72] | 0.77 [0.73, 0.81] | 0.88 [0.86, 0.89] | 0.84 [0.81, 0.87] | 0.91 [0.85, 0.95] | 0.90 [0.84, 0.95] | 0,93 [0.86, 1.00] | 0.91 [0.86, 0.96] | 0,79 [0.70, 0.88] | 0.81 [0.73, 0.89] | 0,75 [0.68, 0.83] | 0.82 [0.75, 0.88] | 0.42 [0.32, 0.53] | 0.24 [0.20, 0.27] | 0.40 [0.35, 0.46] | 0.40 [0.33, 0.46] | 0.69 [0.65, 0.74] | 0.75 [0.69, 0.80] | 0.47 [0.44, 0.50] | 0.47 [0.43, 0.52] |
| Recall (%) | 0.66 [0.54, 0.74] | 0.76 [0.74, 0.77] | 0.64 [0.61, 0.67] | 0.82 [0.78, 0.86] | 0.70 [0.69, 0.72] | 0.75 [0.73, 0.78] | 0.94 [0.92, 0.96] | 0.90 [0.85, 0.94] | 0,19 [0.14, 0.23] | 0.48 [0.42, 0.54] | 0,49 [0.40, 0.58] | 0.63 [0.55, 0.72] | 0,74 [0.66, 0.81] | 0.72 [0.65, 0.79] | 0.25 [0.22, 0.27] | 0.24 [0.21, 0.27] | 0.33 [0.30, 0.38] | 0.31 [0.28, 0.34] | 0.80 [0.74, 0.85] | 0.81 [0.75, 0.86] | 0.54 [0.50, 0.57] | 0.46 [0.42, 0.50] |
| *p* value | <0.05 | | <0.01 | | <0.01 | | 0.33 | | 0.11 | | <0.01 | | 0.08 | | <0.01 | | <0.01 | | <0.01 | | <0.01 | |

### S5 Agreement analysis

**Table S5.1**: Fleiss *k* agreement with 95% CI for each feature extracted (Italian).

| Feature | Llama zero vs Llama few | Llama zero vs Llama few + ann. | Llama zero vs Mistral zero | Llama zero vs Mistral few | Llama zero vs Mistral few + ann. | Llama few vs Mistral zero | Llama few vs Mistral few | Llama few vs Mistral few + ann. | Llama few vs Llama few + ann. | Llama few + ann. vs Mistral zero | Llama few + ann. vs Mistral few | Llama few + ann. vs Mistral | Mistral zero vs Mistral few | Mistral zero vs Mistral few + ann. | Mistral few vs Mistral few + ann. |
|---|---|---|---|---|---|---|---|---|---|---|---|---|---|---|---|

| | | | | | | | | | | | | few + ann. | | | |
|---|---|---|---|---|---|---|---|---|---|---|---|---|---|---|
| Histology | 0.80 [0.75, 0.84] | 0.64 [0.59, 0.69] | 0.78 [0.74, 0.83] | 0.67 [0.62, 0.72] | 0.67 [0.62, 0.72] | 0.75 [0.70, 0.80] | 0.65 [0.60, 0.70] | 0.67 [0.63, 0.72] | 0.63 [0.58, 0.68] | 0.59 [0.54, 0.63] | 0.55 [0.51, 0.60] | 0.74 [0.69, 0.78] | 0.72 [0.67, 0.76] | 0.69 [0.64, 0.75] | 0.64 [0.60, 0.69] |
| Smoking | 0.53 [0.47, 0.58] | 0.64 [0.59, 0.69] | 0.29 [0.23, 0.34] | 0.26 [0.21, 0.31] | 0.31 [0.26, 0.37] | 0.55 [0.50, 0.60] | 0.54 [0.49, 0.60] | 0.50 [0.45, 0.54] | 0.75 [0.70, 0.79] | 0.44 [0.39, 0.49] | 0.44 [0.39, 0.49] | 0.42 [0.36, 0.47] | 0.67 [0.62, 0.72] | 0.51 [0.46, 0.56] | 0.59 [0.54, 0.64] |
| PD-L1 | 0.77 [0.73, 0.81] | 0.77 [0.72, 0.80] | 0.72 [0.68, 0.76] | 0.64 [0.59, 0.68] | 0.53 [0.49, 0.58] | 0.68 [0.63, 0.72] | 0.69 [0.64, 0.73] | 0.59 [0.55, 0.63] | 0.85 [0.82, 0.88] | 0.71 [0.67, 0.75] | 0.74 [0.70, 0.78] | 0.66 [0.61, 0.70] | 0.75 [0.71, 0.79] | 0.65 [0.61, 0.69] | 0.82 [0.78, 0.85] |
| ECOG | 0.92 [0.89, 0.94] | 0.90 [0.88, 0.93] | 0.77 [0.73, 0.81] | 0.76 [0.71, 0.79] | 0.81 [0.77, 0.84] | 0.81 [0.77, 0.82] | 0.78 [0.75, 0.82] | 0.82 [0.769 0.86] | 0.94 [0.92, 0,96] | 0.80 [0.77, 0.84] | 0.78 [0.74, 0.81] | 0.83 [0.79, 0.86] | 0.85 [0.82, 0.88] | 0.86 [0.82, 0.89] | 0.87 [0.84, 0.90] |
| Bone Met | 0.66 [0.60, 0.72] | 0.86 [0.82, 0.90] | 0.65 [0.59, 0.71] | 0.64 [0.58, 0.70] | -0.27 [-0.33, -0.20] | 0.56 [0.48, 0.63] | 0.53 [0.46, 0.60] | -0.47 [-0.53, -0.41] | 0.71 [0.65, 0.77] | 0.65 [0.59, 0.71] | 0.65 [0.59, 0.71] | -0.30 [-0.36, -0.24] | 0.64 [0.58, 0.70] | -0.41 [-0.48, -0.35] | -0.24 [-0.31, -0.17] |
| Liver Met | 0.77 [0.71, 0.84] | 0.84 [0.78, 0.89] | 0.60 [0.52, 0.66] | 0.73 [0.66, 0.79] | 0.73 [0.66, 0.79] | 0.47 [0.39, 0.54] | 0.63 [0.55, 0.70] | 0.58 [0.50, 0.65] | 0.88 [0.83, 0.93] | 0.53 [0.45, 0.60] | 0.66 [0.59, 0.73] | 0.64 [0.56, 0.70] | 0.72 [0.66, 0.77] | 0.72 [0.66, 0.77] | 0.78 [0.73, 0.83] |
| Brain Met | 0.85 [0.80, 0.89] | 0.89 [0.84, 0.93] | 0.77 [0.71, 0.82] | 0.72 [0.65, 0.77] | 0.79 [0.73, 0.84] | 0.69 [0.62, 0.75] | 0.67 [0.61, 0.74] | 0.75 [0.68, 0.81] | 0.92 [0.87, 0.95] | 0.73 [0.67, 0.79] | 0.68 [0.60, 0.73] | 0.76 [0.70, 0.82] | 0.80 [0.75, 0.84] | 0.79 [0.74, 0.84] | 0.78 [0.72, 0.83] |
| T | 0.46 [0.41, 0.51] | 0.15 [0.09, 0.21] | 0.04 [-0.01, 0.08] | 0.01 [-0.03, 0.06] | 0.04 [0.00, 0.08] | 0.11 [0.06, 0.16] | 0.09 [0.05, 0.14] | 0.09 [0.05, 0.14] | 0.03 [-0.04, 0.09] | -0.27 [-0.32, -0.22] | -0.28 [-0.32, -0.23] | -0.24 [-0.28, -0.19] | 0.48 [0.43, 0.53] | 0.40 [0.34, 0.45] | 0.49 [0.44, 0.55] |
| N | 0.65 [0.59, 0.70] | 0.45 [0.38, 0.51] | 0.16 [0.11, 0.22] | 0.22 [0.17, 0.27] | 0.15 [0.10, 0.21] | 0.21 [0.15, 0.26] | 0.24 [0.18, 0.29] | 0.21 [0.15, 0.26] | 0.35 [0.28, 0.41] | -0.01 [-0.06, 0.04] | 0.07 [0.02, 0.12] | -0.04 [-0.09, 0.02] | 0.52 [0.47, 0.56] | 0.45 [0.40, 0.50] | 0.48 [0.43, 0.52] |
| M | 0.45 [0.37, 0.53] | 0.20 [0.12, 0.29] | 0.36 [0.29, 0.43] | 0.34 [0.27, 0.42] | 0.35 [0.28, 0.43] | 0.24 [0.15, 0.31] | 0.23 [0.15, 0.31] | 0.27 [0.19, 0.34] | 0.65 [0.54, 0.74] | 0.08 [0.00, 0.16] | 0.05 [-0.03, 0.13] | 0.13 [0.05, 0.21] | 0.54 [0.48, 0.60] | 0.53 [0.46, 0.60] | 0.57 [0.51, 0.63] |
| Stage | 0.30 [0.22, 0.39] | 0.03 [-0.04, 0.10] | 0.38 [0.31, 0.45] | 0.23 [0.15, 0.30] | 0.09 [0.03, 0.16] | 0.29 [0.21, 0.38] | 0.34 [0.23, 0.43] | 0.26 [0.16, 0.35] | 0.48 [0.35, 0.60] | 0.07 [-0.01, 0.15] | 0.24 [0.13, 0.34] | 0.20 [0.10, 0.31] | 0.39 [0.31, 0.47] | 0.26 [0.18, 0.33] | 0.46 [0.37, 0.55] |
| Median (Q1, Q3) | 0.66 [0.50, 0.79] | 0.64 [0.33, 0.85] | 0.60 [0.32, 0.75] | 0.64 [0.25, 0.70] | 0.35 [0.12, 0.7] | 0.55 [0.27, 0.69] | 0.54 [0.29, 0.66] | 0.50 [0.24, 0.63] | 0.71 [0.55, 0.87] | 0.53 [0.08, 0.68] | 0.55 [0.16, 0.67] | 0.42 [0.05, 0.70] | 0.67 [0.53, 0.74] | 0.53 [0.43, 0.71] | 0.59 [0.49, 0.78] |

**Table S5.2**: Results of Wilcoxon signed-rank test used to compare agreement between two sets of methods and FDR to correct for multiple comparisons.

| Reference | Method A | Method B | Raw p-value | FDR p-value |
|---|---|---|---|---|
| Mistral zero | Llama few | Llama few + ann. | 0.10 | 0.16 |
| Mistral zero | Llama few | Mistral few | 0.001 | 0.009 |
| Mistral zero | Llama few | Llama zero | 0.36 | 0.47 |
| Mistral zero | Llama few | Mistral few + ann. | 0.36 | 0.47 |
| Mistral zero | Llama few + ann. | Mistral few | 0.001 | 0.009 |
| Mistral zero | Llama few + ann. | Llama zero | 0.03 | 0.06 |
| Mistral zero | Llama few + ann. | Mistral few + ann. | 0.08 | 0.14 |
| Mistral zero | Mistral few | Llama zero | 0.01 | 0.04 |
| Mistral zero | Mistral few | Mistral few + ann. | 0.002 | 0.01 |
| Mistral zero | Llama zero | Mistral few + ann. | 0.41 | 0.51 |
| Llama few | Mistral zero | Llama few + ann. | 0.004 | 0.01 |
| Llama few | Mistral zero | Mistral few | 0.76 | 0.80 |
| Llama few | Mistral zero | Llama zero | 0.002 | 0.01 |
| Llama few | Mistral zero | Mistral few + ann. | 0.46 | 0.55 |
| Llama few | Llama few + ann. | Mistral few | 0.004 | 0.01 |
| Llama few | Llama few + ann. | Llama zero | 0.63 | 0.72 |
| Llama few | Llama few + ann. | Mistral few + ann. | 0.004 | 0.014 |
| Llama few | Mistral few | Llama zero | 0.004 | 0.014 |
| Llama few | Mistral few | Mistral few + ann. | 0.24 | 0.36 |
| Llama few | Llama zero | Mistral few + ann. | 0.0009 | 0.009 |
| Llama few + ann. | Mistral zero | Llama few | 0.0009 | 0.009 |
| Llama few + ann. | Mistral zero | Mistral few | 0.83 | 0.85 |
| Llama few + ann. | Mistral zero | Llama zero | 0.001 | 0.009 |
| Llama few + ann. | Mistral zero | Mistral few + ann. | 0.32 | 0.44 |
| Llama few + ann. | Llama few | Mistral few | 0.0009 | 0.009 |
| Llama few + ann. | Llama few | Llama zero | 0.46 | 0.55 |
| Llama few + ann. | Llama few | Mistral few + ann. | 0.001 | 0.009 |

| | | | | |
|---|---|---|---|---|
| Llama few + ann. | Mistral few | Llama zero | 0.01 | 0.04 |
| Llama few + ann. | Mistral few | Mistral few + ann. | 1.0 | 1.0 |
| Llama few + ann. | Llama zero | Mistral few + ann. | 0.03 | 0.06 |
| Mistral few | Mistral zero | Llama few | 0.0009 | 0.009 |
| Mistral few | Mistral zero | Llama few + ann. | 0.002 | 0.01 |
| Mistral few | Mistral zero | Llama zero | 0.002 | 0.01 |
| Mistral few | Mistral zero | Mistral few + ann. | 0.76 | 0.80 |
| Mistral few | Llama few | Llama few + ann. | 0.14 | 0.23 |
| Mistral few | Llama few | Llama zero | 0.89 | 0.91 |
| Mistral few | Llama few | Mistral few + ann. | 0.06 | 0.12 |
| Mistral few | Llama few + ann. | Llama zero | 0.32 | 0.44 |
| Mistral few | Llama few + ann. | Mistral few + ann. | 0.05 | 0.10 |
| Mistral few | Llama zero | Mistral few + ann. | 0.06 | 0.12 |
| Llama zero | Mistral zero | Llama few | 0.006 | 0.01 |
| Llama zero | Mistral zero | Llama few + ann. | 0.36 | 0.47 |
| Llama zero | Mistral zero | Mistral few | 0.17 | 0.26 |
| Llama zero | Mistral zero | Mistral few + ann. | 0.41 | 0.51 |
| Llama zero | Llama few | Llama few + ann. | 0.27 | 0.40 |
| Llama zero | Llama few | Mistral few | 0.0009 | 0.009 |
| Llama zero | Llama few | Mistral few + ann. | 0.0009 | 0.009 |
| Llama zero | Llama few + ann. | Mistral few | 0.10 | 0.16 |
| Llama zero | Llama few + ann. | Mistral few + ann. | 0.04 | 0.08 |
| Llama zero | Mistral few | Mistral few + ann. | 0.70 | 0.77 |
| Mistral few + ann. | Mistral zero | Llama few | 0.001 | 0.009 |
| Mistral few + ann. | Mistral zero | Llama few + ann. | 0.08 | 0.14 |
| Mistral few + ann. | Mistral zero | Mistral few | 0.01 | 0.03 |
| Mistral few + ann. | Mistral zero | Llama zero | 0.02 | 0.05 |
| Mistral few + ann. | Llama few | Llama few + ann. | 0.57 | 0.66 |
| Mistral few + ann. | Llama few | Mistral few | 0.001 | 0.009 |
| Mistral few + ann. | Llama few | Llama zero | 0.76 | 0.80 |
| Mistral few + ann. | Llama few + ann. | Mistral few | 0.006 | 0.01 |
| Mistral few + ann. | Llama few + ann. | Llama zero | 0.57 | 0.66 |

| | | | | |
|---|---|---|---|---|
| Mistral few + ann. | Mistral few | Llama zero | 0.006 | 0.01 |

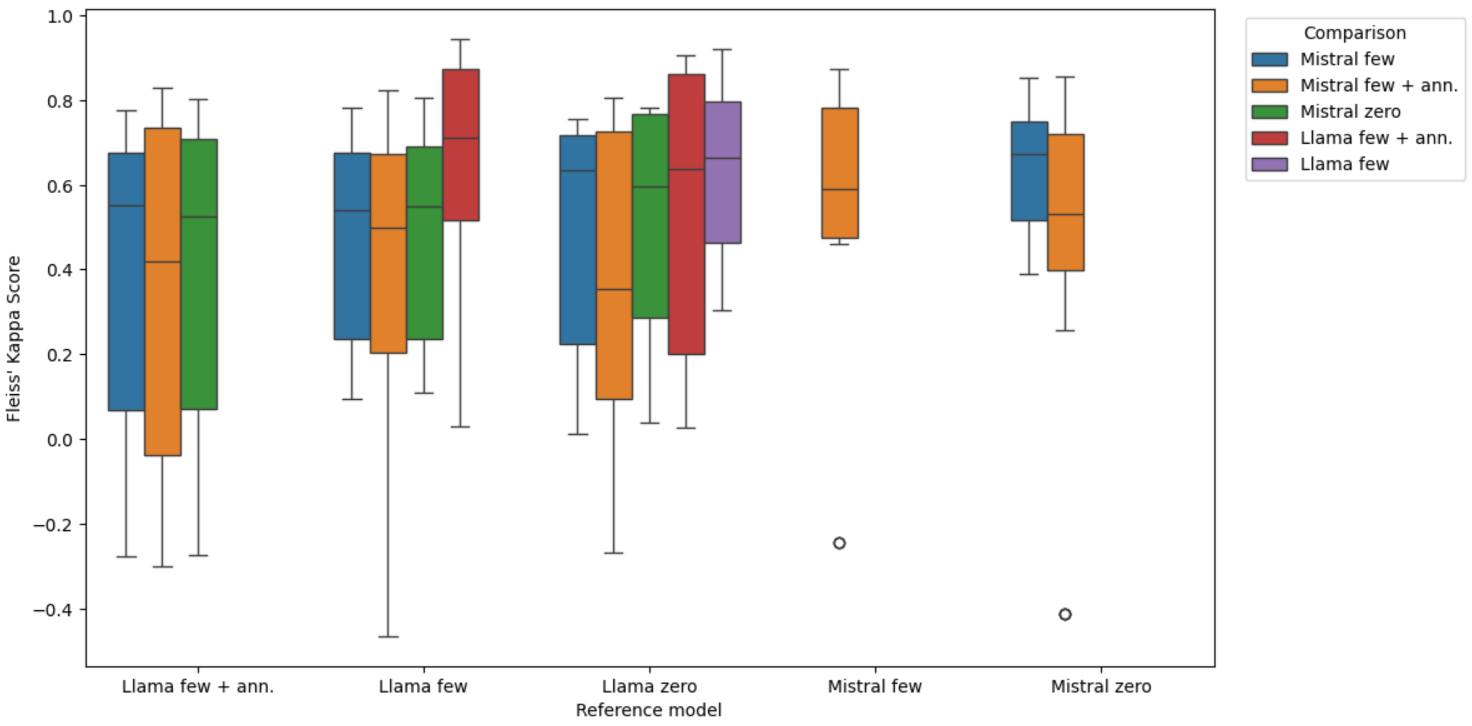


Figure S5.3 Boxplot showing the pairwise agreement of LLMs (LLaMa 3.1 8B and Mistral) with different Italian prompting strategies. Each box represents the distribution of Fleiss' Kappa values calculated across all cancer-related features for each model-prompt pair.

### S6 Missing information analysis

***Missing information analysis***

As additional analysis, we investigated the hallucination rate associated with the inference of MI (i.e., the “non mentionated” label), under both zero-shot and few-shot+ann prompting. For instance, under zero-shot conditions (Table S6.1) we observed that, for smoking and ECOG PS with LLaMa 3.1, the hallucination rate is 48.09% and 32.84%, respectively (vs 33.5% and 9.42%in the GT). With Mistral and smoking and ECOG PS, the hallucination rate decreases to 9.79% and 13.43%, proving that this model tends to hallucinate less towards the “not mentionated” label. However, under a few-shot+ann design (Table S6.2), the rate of hallucination reduces to18.05.% and 19.4% with LLaMa 3.1, respectively. This analysis demonstrates that the inference of real missing information is improved with few-shot + ann prompting. Finally, in supplementary Figures S6.3 and S6.4 we report examples of MI inference for 5 patients’ cases in which PD-L1<1% has been wrongly inferred as “not mentioned”. These examples align with the main analysis proving better extraction with few-shot+ann.

Table S6.1: Analysis of missing value information in zero-shot prompting.

| | | **Llama 3.1 8B** | | **Mistral 7B** | |
|---|---|---|---|---|---|
| **Clinical Features** | MI (GT). (n/%) | Number of matches | Halluc. (n/%) | Number of matches | Halluc. (n/%) |
| **Histology** | 6 (0.84%) | 6 | 0 (0.00%) | 6 | 0 (0.00%) |
| **Smoking** | 235 (33.5%) | 122 | 113 (48.09%) | 212 | 23 (9.79%) |
| **PD-L1** | 205 (28.83%) | 193 | 7 (3.41%) | 184 | 14 (6.83%) |
| **ECOG PS** | 67 (9.42%) | 45 | 22 (32.84%) | 58 | 9 (13.43%) |
| **T (TNM)** | 5 (0.70%) | 0 | 5 (100%) | 1 | 4 (100%) |
| **N (TNM)** | 4 (0.56%) | 0 | 4 (100%) | 1 | 3 (100%) |
| **Stage** | 5 (0.70%) | 0 | 5 (100%) | 0 | 5 (100%) |

Table S6.2: Analysis of missing value information in few-shot +ann data.

| | | Llama 3.1 8B | | Mistral 7B | |
|---|---|---|---|---|---|
| **Clinical Features** | MI (GT). (n/%) | Number of matches | Halluc. (n/%) | Number of matches | Halluc. (n/%) |
| **Histology** | 6 (0.84%) | 5 | 1 (16.67%) | 4 | 2 (33.33%) |
| **Smoking** | 235 (33.5%) | 198 | 37 (18.05%) | 138 | 97 (41.28%) |
| **PD-L1** | 205 (28.83%) | 193 | 7 (3.41%) | 148 | 52 (25.37%) |
| **ECOG PS** | 67 (9.42%) | 54 | 13 (19.40%) | 52 | 14 (20.90%) |
| **T (TNM)** | 5 (0.74%) | 0 | 5 (100%) | 1 | 5 (100%) |
| **N (TNM)** | 4 (0.56%) | 0 | 4 (100%) | 0 | 4 (100%) |
| **Stage** | 5 (0.74%) | 0 | 5 (100%) | 1 | 4(100%) |

### S7 Medical expertise experiment

**Table S7.1** Fleiss *k* agreement with 95% CI for each feature extracted in the zero-shot phase of the experiment.

| Feature | Experienced | Resident | Student | Experienced vs student | Resident vs Student | Experienced vs Resident |
|---|---|---|---|---|---|---|
| Bone Met | 0.79 [0.79, 0.80] | 0.71 [0.71, 0.72] | 0.54 [0.54, 0.55] | 0.67 [0.67, 0.68] | 0.63 [0.63, 0.64] | 0.75 [0.74, 0.75] |
| Brain Met | 0.88 [0.87, 0.88] | 0,75 [0.74, 0.75] | 0.88 [0.88, 0.89] | 0.88 [0.87, 0.88] | 0.81 [0.81, 0.82] | 0.82 [0.81, 0.82] |
| Liver Met | 0.82 [0.81, 0.82] | 0.70 [0.69, 0.70] | 0.70 [0.69, 0.70] | 0.76 [0.75, 0.76] | 0.69 [0.68, 0.69] | 0.76 [0.75, 0.76] |
| T | 0.18 [0.18, 0.19] | 0.25 [0.25, 0.26] | 0.32 [0.31, 0.32] | 0.24 [0.24, 0.25] | 0.30 [0.29, 0.30] | 0.23 [0.22, 0.23] |

| | | | | | | |
|---|---|---|---|---|---|---|
| N | 0.35 [0.35, 0.36] | 0.34 [0.33, 0.35] | 0.40 [0.39, 0.41] | 0.37 [0.37, 0.38] | 0.38 [0.37, 0.38] | 0.36 [0.35, 0.36] |
| M | 0.44 [0.44, 0.45] | 0.21 [0.21, 0.22] | 0.37 [0.37, 0.38] | 0.41 [0.40, 0.41] | 0.32 [0.31, 0.32] | 0.34 [0.34, 0.35] |
| Stage | 0.28 [0.28, 0.29] | 0.22 [0.22, 0.23] | 0.46 [0.45, 0.46] | 0.36 [0.35, 0.36] | 0.33 [0.33, 0.34] | 0.33 [0.33, 0.34] |
| Median (Q1, Q3) | 0.44 [0.32, 0.81] | 0.34 [0.24, 0.7] | 0.46 [0.38, 0.62] | 0.41[0.36, 0.72] | 0.38 [0.32, 0.66] | 0.36 [0.34, 0.76] |

**Table S7.2** Fleiss *k* agreement with 95% CI for each feature extracted in the few-shot phase of the experiment.

| Feature | Experienced | Resident | Student | Experienced vs student | Resident vs Student | Experienced vs Resident |
|---|---|---|---|---|---|---|
| Bone Met | 0.79 [0.79, 0.80] | 0.53 [0.53, 0.54] | 0.64 [0.63, 0.64] | 0.71 [0.70, 0.71] | 0.57 [0.56, 0.57] | 0.68 [0.67, 0.68] |
| Brain Met | 0.79 [0.79, 0.80] | 0.47 [0.46, 0.47] | 0.77 [0.77, 0.77] | 0.78 [0.77, 0.78] | 0.61 [0.60, 0.61] | 0.64 [0.64, 0.64] |
| Liver Met | 0.90 [0.90, 0.90] | 0.67 [0.66, 0.67] | 0.86 [0.85, 0.86] | 0.87 [0.87, 0.88] | 0.76 [0.76, 0.76] | 0.80 [0.80, 0.80] |
| T | 0.35 [0.35, 0.36] | 0.37 [0.36, 0.38] | 0.51 [0.50, 0.51] | 0.43 [0.42, 0.43] | 0.45 [0.44, 0.45] | 0.37 [0.37, 0.37] |
| N | 0.24 [0.23, 0.24] | 0.35 [0.34, 0.36] | 0.45 [0.44, 0.46] | 0.33 [0.33, 0.34] | 0.41 [0.41, 0.42] | 0.30 [0.29, 0.30] |
| M | 0.17 [0.16, 0.17] | 0.22 [0.21, 0.22] | 0.50 [0.49, 0.50] | 0.29 [0.28, 0.29] | 0.35 [0.34, 0.35] | 0.22 [0.21, 0.22] |
| Stage | 0.08 [0.08, 0.09] | 0.21 [0.20, 0.22] | 0.38 [0.37, 0.38] | 0.21 [0.20, 0.21] | 0.30 [0.29, 0.30] | 0.16 [0.16, 0.16] |
| Median (Q1, Q3) | 0.35 [0.20, 0.79] | 0.37 [0.29, 0.50] | 0.51 [0.48, 0.70] | 0.43 [0.31, 0.74] | 0.45 [0.38, 0.59] | 0.37 [0.26, 0.66] |

**Table S7.3** Results of Wilcoxon rank test used to compare the intra and inter-rater Fleiss's *k* agreement between the two phases of the experiments.

| Group | Wilcoxon p-value |
|---|---|
| experienced | 0.08 |
| resident | 0.29 |

| | |
|---|---|
| student | 0.03 |
| experienced vs student | 0.46 |
| resident vs student | 0.81 |
| experienced vs resident | 0.04 |

### S8 TKI cohort results

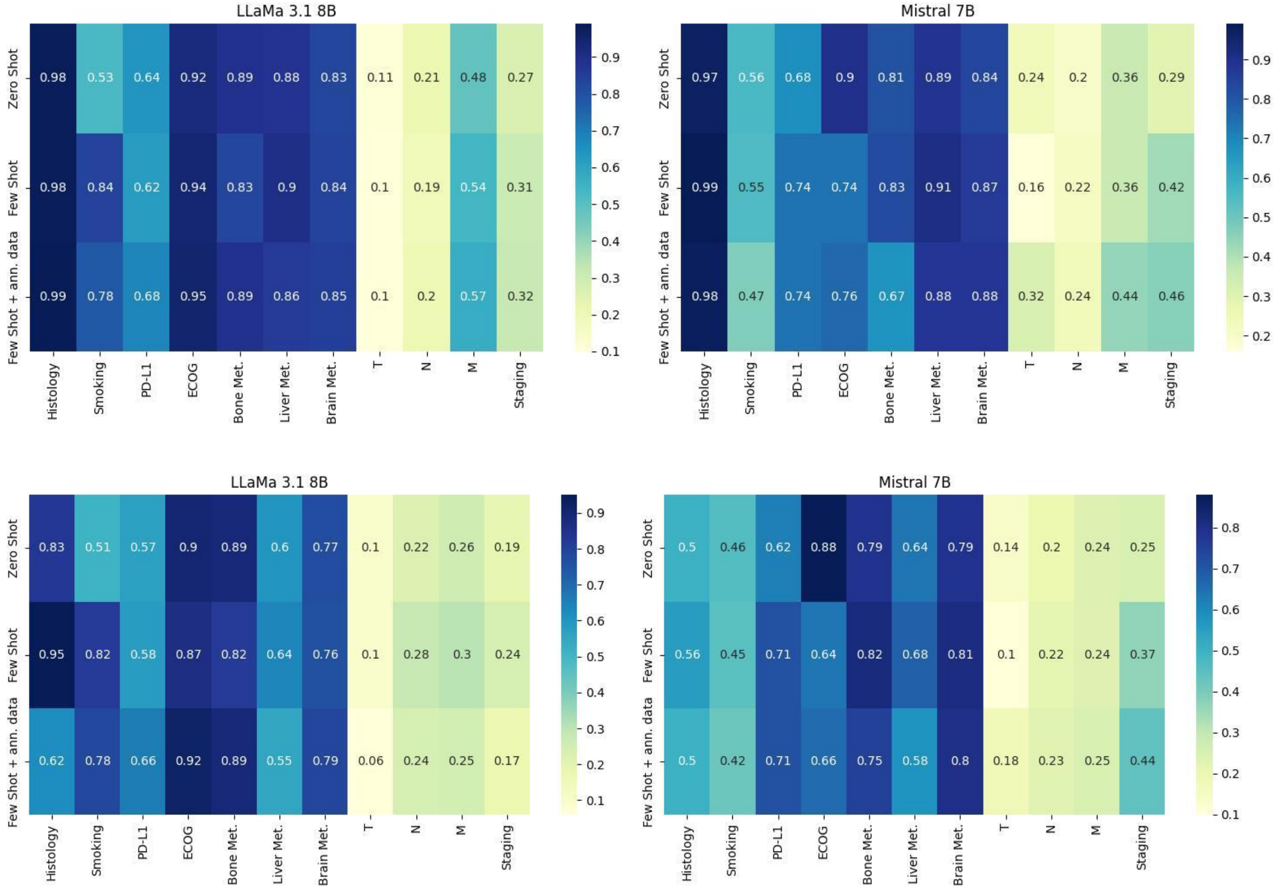


Figure S8.1: Heatmaps of a) accuracy and b) F1-Score for LLaMa 3.1 8B and Mistral 7B by prompting technique on the TKI cohort.
